# Counterfactual evaluation of elementary and secondary school policies in the COVID-19 pandemic

**DOI:** 10.1101/2025.08.17.25333861

**Authors:** Benedetta Canfora, Rey Audie Escosio, Otilia Boldea, Ana Nunes, Ganna Rozhnova

## Abstract

Understanding the role of elementary and secondary schools in SARS-CoV-2 transmission is essential for designing effective public health policies. We developed a stochastic age-stratified transmission model, fitted to Dutch epidemiological and contact survey data, to evaluate how alternative closure and reopening strategies for elementary and secondary schools might have shaped the course of the pandemic in the Netherlands from early 2020 through late 2021. Using counterfactual simulations combined with time-varying elasticity analysis, we found that school closures had the potential to mitigate transmission and reduce burden on healthcare. However, their effectiveness was highly context-dependent, influenced by school type, contact patterns, policy timing, and population immunity. Our findings indicate that the contributions of different age groups to transmission shifted over time. Adults were the primary drivers early in the pandemic, followed by adolescents in late 2020 and early 2021, and children in late 2021. These dynamics were reflected in the changing relative impact of different educational levels on overall transmission and hospital burden. Secondary schools had a greater effect on national hospitalizations in 2020, while elementary schools became more important in 2021, partly due to lower prior infection-induced immunity and negligible vaccination coverage among children compared to adolescents. Together, these results highlight the importance of accounting for age-specific transmission dynamics and changing epidemiological landscape when evaluating school closure and reopening strategies. Our study supports a more targeted and adaptive approach to school policies in future pandemics.

## Introduction

The role of schools in SARS-CoV-2 transmission has been a subject of scientific and policy debate throughout the COVID-19 pandemic [1–6]. Early responses in many countries included widespread school closures, based on historical evidence from influenza outbreaks where limiting school-based contacts helped reduce transmission [7, 8]. As the pandemic progressed, decisions on school closures and reopenings were repeatedly revised in response to changing epidemiology, emerging variants, and rising pressure on healthcare systems. Yet, despite their widespread use throughout the pandemic, the population-level impact of school closures and reopenings differentiated by educational level remains incompletely understood.

This gap is critical given the age-dependent nature of SARS-CoV-2 epidemiology. Disease severity and susceptibility to infection increase with age, with younger children estimated to be less susceptible and to experience many asymptomatic infections [4, 9–17]. Contact patterns in schools also differ by educational level, with adolescents typically engaging in more transmission-relevant interactions than younger children [18–21]. In addition, vaccination eligibility, uptake, and timing varied across age groups, with younger children often the last to become eligible. These biological and behavioral differences, combined with the considerable social and educational costs of school closures [22–27], highlight the importance of distinguishing between elementary and secondary schools when designing or evaluating school policies.

Two key considerations emerge in the context of counterfactual evaluation of historical school policies. One is whether keeping schools open during periods of closure could have been a viable option without triggering major waves of hospitalizations. The other is whether closing schools during periods of widespread societal restrictions, when they remained open, could have played a decisive role in preventing or reducing hospitalization peaks.

To date, most modeling studies on school interventions have focused on forward projections rather than retrospective evaluations of what might have happened under alternative policy choices [28–35]. Counterfactual analyses of school closures and reopenings are scarce and limited in scope [36–38]. Specifically, they concern the early phase of the pandemic [36, 37], do not quantify the distinct contributions of educational levels [36, 38], and focus only on school openings without assessing alternative school closures [37, 38]. Many observational and within-school modeling studies aimed to understand transmission and interventions in educational settings or school-aged populations (e.g., [39–49]). However, these approaches are usually not suitable for measuring the impact of elementary and secondary schools on population-level hospitalizations or conducting counterfactual analyses. A handful of studies used elasticity analyses to reconstruct actual age-specific transmission dynamics at national scales, but they did not include counterfactual scenarios of school interventions [50–52]. To our knowledge, no previous modeling study has systematically and comprehensively evaluated how school closure and reopening strategies targeted at different educational levels may have shaped the course of the pandemic during its critical first two years.

Here, we present one of the most detailed retrospective modeling analyses of elementary and secondary school policies to date. As a case study, we focus on the Netherlands, where pandemic school strategies shifted over time, alternating between full closures, partial openings, and full reopenings. We use an age-structured transmission model to evaluate the impact of school closures during full lockdowns and school reopenings during semi-lockdowns across four periods in 2020–2021, each marked by major changes in restrictions and viral dynamics. Our counterfactual analysis distinguishes between elementary and secondary schools, allowing us to isolate their respective contributions to population-level infections and hospitalizations. We further quantify the age-specific contributions to transmission over time using elasticity analysis, offering a dynamic view of transmission drivers. These analyses provide policy-relevant insights into the role of different educational levels in virus spread, with implications for future pandemic preparedness.

## Methods

### Overview

We extended a stochastic, age- and region-stratified compartmental transmission model, fitted to regional reported cases, hospitalizations, national contact surveys, vaccination, and seroprevalence data, developed in [53]. Parameter estimation was performed using a quasi-Bayesian approach based on the ensemble adjustment Kalman filter. Compared to the previous model, our study differed in how social contacts were modeled, using an extensive set of contact survey data. The model was implemented in MATLAB (version R2024a).

### Data

The model was calibrated using demographic, epidemiological, mobility, vaccination, and variant-related data from the Netherlands. Demographic data stratified by age and province were obtained from the Dutch Central Bureau of Statistics [54]. Daily reported PCR-confirmed infections by age and province, as well as multiple hospitalization datasets, covering different periods and levels of aggregation, were obtained from the National Institute for Public Health and the Environment (RIVM). National seroprevalence data were derived from four rounds of the PIENTER Corona Study, coordinated by the RIVM [55]. Mobility patterns were reconstructed using train travel data from the Dutch Railways from the start of the pandemic until September 2021, extended with Google mobility data through January 2022 [56]. Vaccination coverage, including boosters, and genomic surveillance data on variant circulation were obtained from the official RIVM sources [57]. Except for contact data, which have been updated using newly available surveys, all data sources are consistent with those used in [53], to which we refer the reader for further details.

### Transmission model

This work builds upon the model presented in [53]. The developed model is a stochastic compartmental model stratified by age, region, and disease status. The population is categorized into children (0 to 9 years old), adolescents (10 to 19 years old), and adults (above 19 years old) across twelve Dutch provinces. This population stratification is further complemented by real-world mobility for the Dutch population. To align with Dutch government recommendations advising infected individuals to stay at home, the model assumes that individuals with undocumented infections may travel and contribute to transmission between provinces, while those with documented infections contribute to transmission only within their province of residence.

A schematic representation of the model is provided in Figure 1. The model differentiates between individuals with no prior exposure and those who have previously been infected or vaccinated. Individuals in the fully susceptible class (*S*^1^) become latently infected (*E*^1^) with an age-specific force of infection, *β*^1^. After an average latent period *Z*, they become infectious (*I*^1^). An age-specific proportion *α* of these cases is detected and reported (*I*^*R*,1^), while the remaining proportion (1−*α*) is unreported (*I*^*U*,1^). Both groups recover without hospitalization (*R*^1^) after an average infectious period *D*. Only individuals who have a reported infection become hospitalized (*H*^1^) at an age-specific rate *γ*^1^. Hospitalized individuals recover after an average duration of 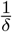, which differs by age group. Upon recovery, immunity wanes at a rate *η*^1^, and individuals transition to a partially susceptible state (*S*^2^). These individuals can be reinfected, now under a modified force of infection *β*^2^, and follow the same disease progression as primary infections. Immunity after reinfection also wanes (at rate *η*^2^), returning individuals to *S*^2^. Vaccination occurs at an age-specific rate *ν* and is targeted to all individuals except those currently hospitalized or with a reported primary infection. For parsimony, individuals who are either vaccinated and susceptible or have lost immunity after recovering from a primary infection are grouped into a single partially susceptible class (*S*^2^). Reinfections and breakthrough infections (i.e., infections occurring after vaccination) are also grouped under this structure. Both vaccination and prior infection influence transmission in three ways: they reduce susceptibility (affecting *β*^2^), infectiousness (not depicted), and the probability of hospitalization upon reinfection or breakthrough infection (modeled via *γ*^2^). The population size is assumed to remain constant throughout the study period. The model equations are provided in the Supplementary Material, Section S1.

**Figure 1.**
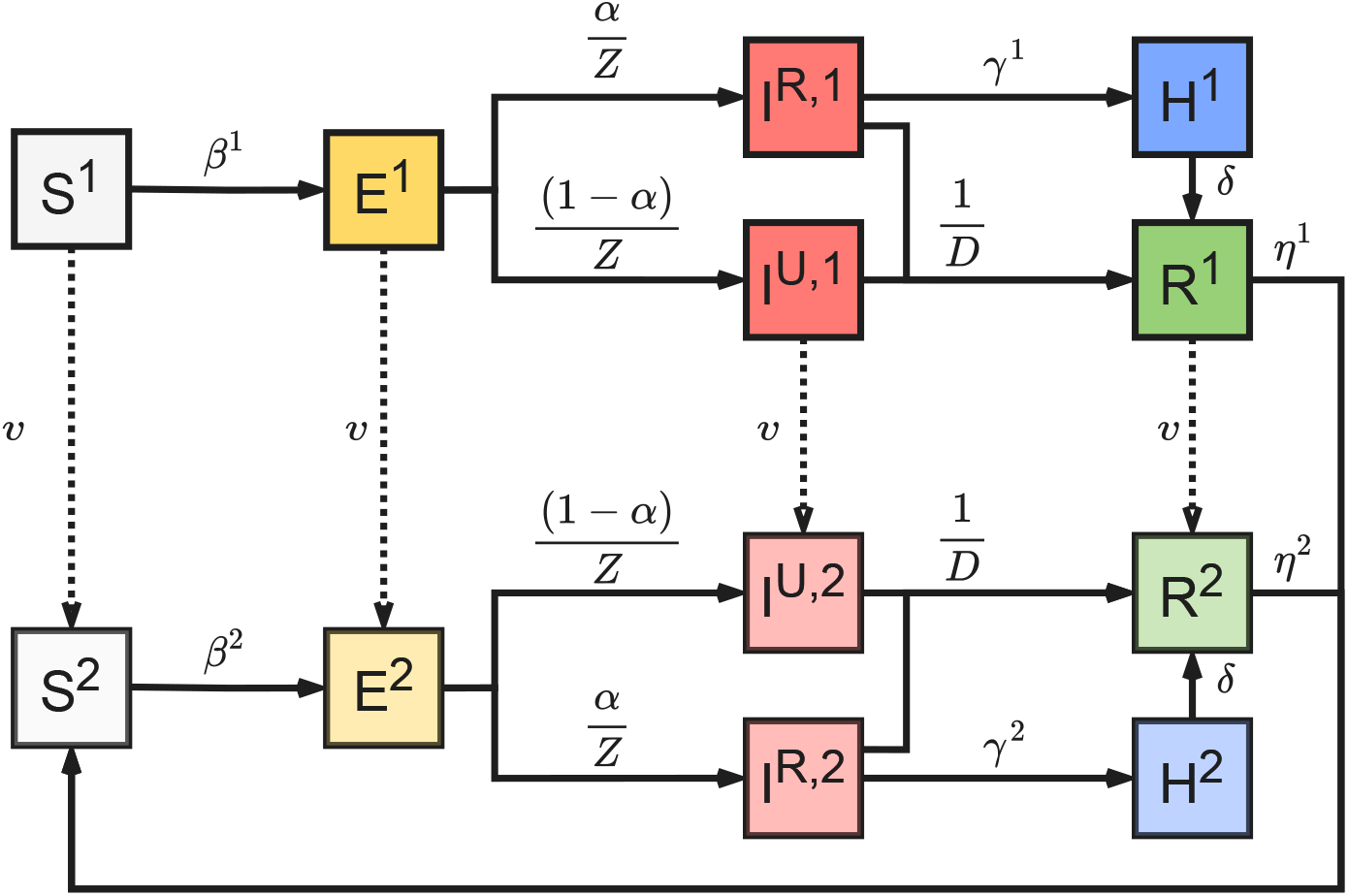
Schematic of the transmission model. Black arrows show epidemiological transitions. Dashed arrows indicate vaccination. The top set of compartments denotes individuals who are unvaccinated or who have not yet lost immunity after primary infection. The bottom set of compartments denotes individuals who are vaccinated or those who have lost immunity after primary infection. The age-specific parameters were forces of infection, *β*^1^ and *β*^2^, case detection rate, *α*, hospitalization rates, *γ*^1^ and *γ*^2^, duration of hospitalization, 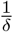, and vaccination rate, *ν*. The remaining parameters did not depend on age. For simplicity, the movement of individuals between provinces is not shown.

### Contact patterns

Contact patterns were modeled using 3 × 3 matrices that represent the average daily number of close contacts between children, adolescents, and adults. Following the contact structure in Boldea et al. [53], which builds upon Backer et al. [58], we stratify total contacts into school and non-school components, further disaggregating school contacts into elementary school (E) and secondary school (S) levels. In the original model, school contacts, when schools were open, were assumed to remain constant over time and were based on pre-pandemic estimates, while non-school contacts varied dynamically across several behavioral regimes corresponding to key phases of the pandemic.

In this study, while we retain the conceptual distinction between school and non-school contacts and the E/S split, we adopt a different approach. Due to a newly available extensive set of contact surveys, covering a broad range of periods, we base our modeling on observed total contacts and then partition them into school and non-school components. For each survey, school contacts are estimated to be a fraction 0 ≤ *q* ≤ 1 of the pre-pandemic school contacts [53,58]. The residual contacts, after accounting for school interactions, are attributed to non-school settings. The fraction *q*, specific to each survey, is estimated in such a way as to satisfy two constraints: (i) non-school contacts must remain non-negative, and (ii) they must not fall below the level observed during the first lockdown, which serves as a lower bound due to the strict containment measures at that time. This approach retains the separation between elementary (E) and secondary (S) school contacts, assuming additivity because of the physical separation of Dutch elementary and secondary schools, but it takes into account that even when schools were open at full capacity, contacts may be lower in schools due to preventive measures. Since survey data are not available for all periods, we performed grid searches to infer contact patterns during the missing intervals. School contact patterns may vary across provinces due to the staggered opening and closing of schools between the northern, central, and southern regions of the Netherlands. Further details of available contact surveys, contact matrices, and contact modeling are provided in Supplementary Table S1, Supplementary Figures S1-S4, and Supplementary Material, Section S2.

### Parameter estimation

The fitting method was a quasi-Bayesian estimation based on the ensemble adjustment Kalman filter, which efficiently integrates various data sources. Filtering techniques were used to estimate the likelihood of observed data and to update system states when some variables are unobserved. Originally proposed by [59], the ensemble adjustment Kalman filter is a variant of the ensemble Kalman filter suitable for high-dimensional, stochastic models. In our implementation, a large number of ensemble members are drawn from prior distributions over all parameters and state variables. The ensemble of draws evolves according to a stochastic model that captures age- and region-specific dynamics and the stochasticity needed due to smaller populations in age-region groups. Model updates are performed sequentially using provincial and neighboring region data, allowing efficient assimilation of information for a large number of states. Compared to particle filters and related methods, which often suffer from degeneracy in high dimensions due to resampling [60, 61], this method maintains diversity of draws without resampling. It thus offers a compromise between traditional Kalman filters requiring linearity and normality and more flexible but less computationally stable alternatives.

#### Initialization and parameter identifiability

Model parameter estimation largely followed the approach described in [53]. Previous parameter calibrations were retained. To ensure identifiability, additional parameters were calibrated, using estimates from [53] and other relevant studies. In particular, the initial number of infected individuals was derived from [28]. The posterior distributions were examined, verifying that they became sufficiently concentrated around specific values to signal learning from the data. The estimated parameters vary across the periods considered, reflecting the separate model fitting for each period, where the posterior distributions from one period were used to initialize the next period. For each period, we identified the subset of parameters to be estimated and performed grid searches to fix the remaining parameters, ensuring that the baseline counterfactual, based on simulations with the estimated model parameters, closely matched the real data. This step was essential for internal model validity, given the variations in data and epidemic context over time. Further details of the model parameters, parameter posterior distributions, the model fit and baseline counterfactual are provided in Supplementary Table S2 and Supplementary Figures S5-S9.

### Counterfactual periods and scenarios

We conducted counterfactual analyses in four periods (CP1, CP2, CP3, and CP4), each characterized by different combinations of the observed school status and broader societal restrictions, ranging from full lockdowns (LD1 and LD2) to semi-lockdowns (SLD1 and SLD2) (Figure 2). For each period, we compared a baseline scenario that reproduces observed data with three counterfactual scenarios designed to assess the impact of reverting the observed school status. In CP1 and CP3, when schools were closed, we simulated the impact of reopening: (i) both elementary and secondary schools, (ii) only elementary schools, and (iii) only secondary schools. Conversely, in CP2 and CP4, when schools remained open, we evaluated the impact of closing them under the same three configurations. The counterfactual scenarios differed from the baseline only in terms of school-related contacts. All other epidemiological parameters and non-school interventions remained unchanged.

**Figure 2.**
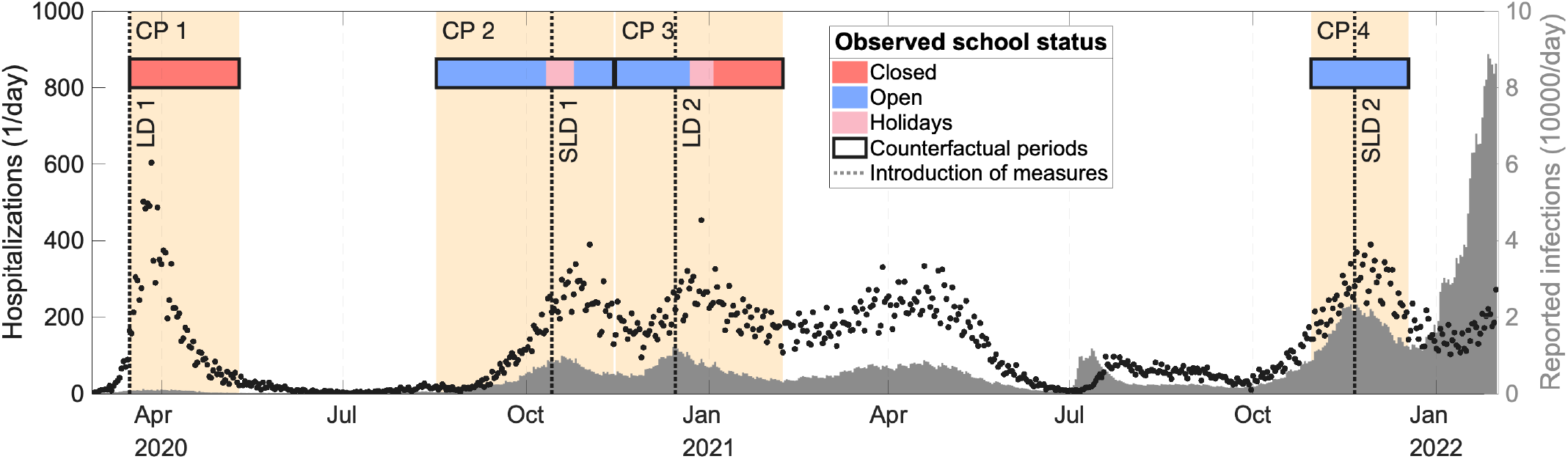
Schematic timeline of the pandemic in the Netherlands. Black dots represent daily hospital admissions (left axis), and the gray area shows daily reported infections (right axis), spanning from early 2020 to early 2022. Shaded areas highlight the counterfactual periods—CP1, CP2, CP3, and CP4—which are defined around the true date of specific measures implementation (full lockdowns, LD, and semi-lockdowns, SLD). The timing of these measures is indicated by vertical dashed gray lines. The observed school status is shown with colored bars: red for closed, blue for open, and pink for holidays. Black-outlined segments mark the periods used for counterfactual analyses. During CP1 schools were closed, and we evaluated the hypothetical impact of maintaining them open. During CP2 and CP4 schools were open, and we evaluated the hypothetical impact of closing them. During CP3 schools were closed for only part of the time, and we assessed the potential effect of keeping them open in that subperiod.

The four periods were selected around the implementation of major lockdowns and semi-lockdowns:

1. CP1 (16 March – 10 May 2020): Both elementary and secondary schools were closed during the first lockdown (LD 1) that began on 16 March. We evaluated the impact of reopening schools over the entire period.
2. CP2 (17 August – 14 November 2020): Both elementary and secondary schools were open, except for one-week autumn holidays with timing varied by province. A semi-lockdown (SLD 1) was introduced in October. We assessed the impact of closing schools over the entire period.
3. CP3 (15 November 2020 – 7 February 2021): The second lockdown (LD 2) was introduced in December. Elementary schools remained closed after the Christmas holidays, while secondary schools reopened at reduced capacity (25%). We evaluated the impact of reopening schools to full capacity after the Christmas holidays.
4. CP4 (30 October – 18 December 2021): Both elementary and secondary schools remained open during a period of gradually enacted semi-lockdown (SLD 2). We assessed the impact of closing schools over the entire period.

### Reproduction number and elasticity analysis

To quantify the transmission potential of the virus, we use the effective reproduction number, ℛ_*e*_(*t*). For each time *t*, the value of ℛ_*e*_(*t*) is obtained as the largest eigenvalue of the next-generation matrix. However, obtaining the next-generation matrix using the current model is computationally expensive due to stratifications by age and region, stochasticity, and the long period over which the study is conducted. Since epidemiological parameters do not vary across regions, we approximate ℛ_*e*_(*t*) using a simplified model that accounts only for age (adults, adolescents, and children) and immunity status (naïve and partially immune after vaccination or prior infection). The full derivation of the next-generation matrix using techniques described in [62, 63] is provided in the Supplementary Material, Section S5.

Elasticity matrices, ℰ_*mn*_(*t*), are constructed using the next-generation matrices and the effective reproduction number. Here, indices *m* and *n* each refer to one of six population groups defined by the combination of age and immunity status. From these matrices, we derive the cumulative elasticity for each group *m* as *e*_*m*_(*t*) = ∑_*n*_ ℰ_*mn*_(*t*). Each cumulative elasticity, *e*_*m*_(*t*), quantifies the time-varying contribution of group *m* to the effective reproduction number, ℛ_*e*_(*t*). The derivation of *e*_*m*_(*t*) can be found in the Supplementary Material, Section S6.

### Model outcome measures

After fitting the model for each of the four periods to observed infections and hospitalizations, we used it to generate the corresponding baseline counterfactual scenarios (Supplementary Figure S9). For each period and counterfactual scenario, we computed (i) the relative change in cumulative daily infections and hospitalizations relative to the baseline, by age group and for the total population; (ii) bed occupancy over time, to evaluate how changes in measures affect hospital burden; and (iii) the effective reproduction number and cumulative elasticities to demonstrate how each age group contributes to transmission. All outcomes are reported as medians, except for cumulative elasticities and contacts, which are reported as means. Uncertainty in the model outcomes is reported as the 95% posterior credible intervals based on 300 posterior updates. Additional model outcomes are provided in Supplementary Materials, Sections S7-S9.

## Results

### First counterfactual period: CP1

In CP1, which includes the first lockdown, we assessed the impact of keeping schools open during a period when they were, in reality, closed (Figure 3). Maintaining schools open in CP1 would have led to an increase in infections across all age groups, with particularly pronounced surges among children and adolescents (Figure 3**A**). In the worst-case scenario, where both elementary and secondary schools remained open, infections in adolescents and children would have increased by more than 2400% (95% CrI 1184%–3728%) and more than 1100% (95% CrI 612%–3245%), respectively, compared to the baseline scenario. This increase in infections among children and adolescents would have amplified onward transmission to adults, ultimately resulting in higher numbers of adult hospitalizations, even though children and adolescents themselves rarely require hospitalization. Cumulative hospitalizations across the total population would have increased by 144% (95% CrI 109%–221%) — a rise driven almost entirely by hospitalizations in adults. A similar pattern is observed across all CPs: changes in total hospitalizations, whether increases following school openings or reductions following closures, are consistently dominated by adult admissions, while hospitalizations among children remain much lower throughout.

**Figure 3.**
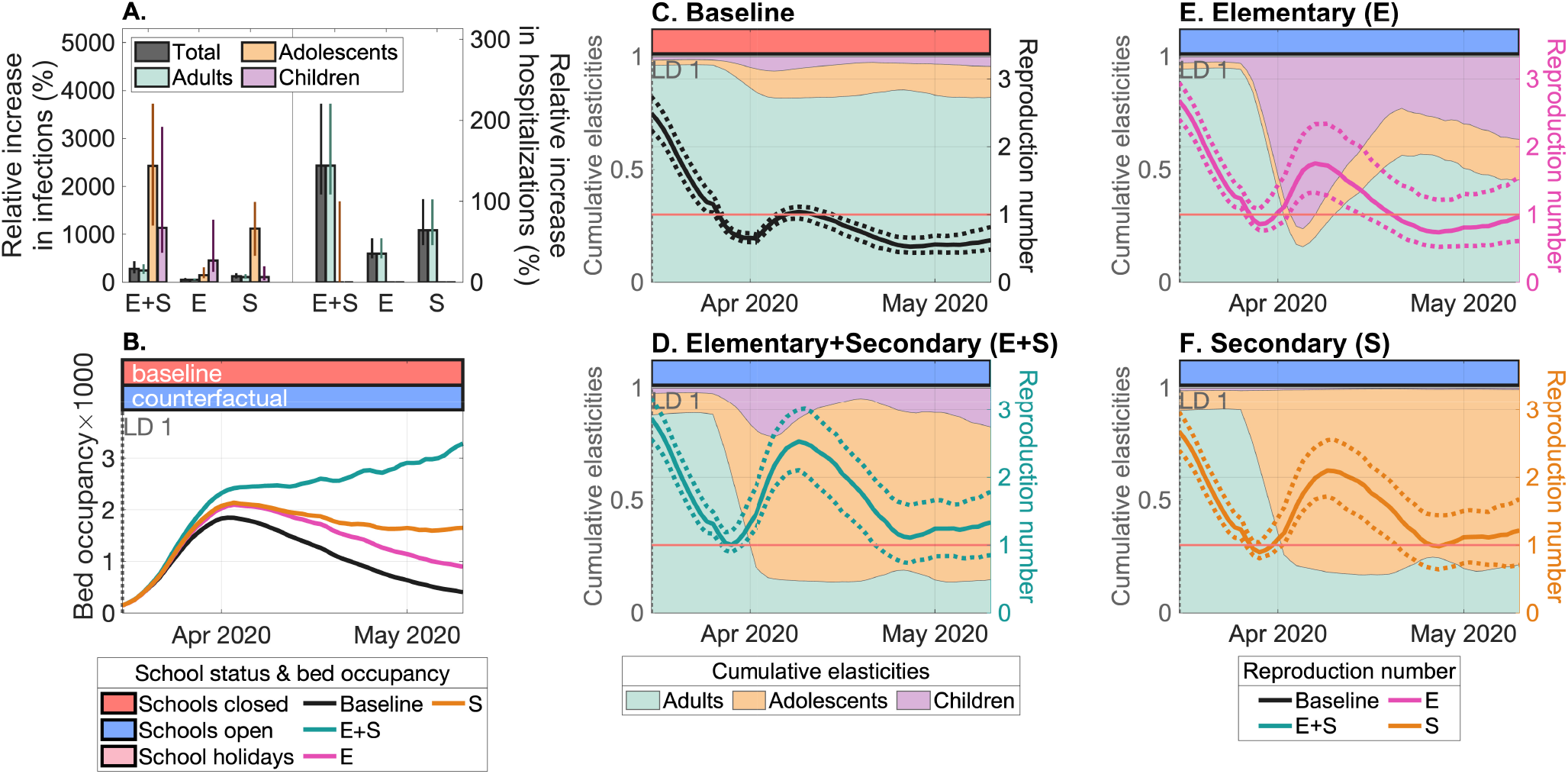
Retrospective evaluation of opening elementary and secondary schools in CP1. The colored horizontal bars in (**B**)-(**F**) indicate the status of school operations in the baseline and counterfactual scenarios. **(A)** Relative increase in cumulative infections and hospitalizations in the counterfactual scenarios compared to the baseline, by age group and for the total population. The bars represent median values, and the whiskers indicate the 95% posterior credible intervals. **(B)** Projected median hospital bed occupancy over time for the baseline and counterfactual scenarios. (**C**)-(**F**) Mean cumulative elasticities by age group (left y-axis) and median effective reproduction number (right y-axis) for **(C)** the baseline and three counterfactual scenarios: **(D)** full school opening, **(E)** opening of elementary schools only, and **(F)** opening of secondary schools only. In (**C**)-(**F**) the shaded areas represent the contributions of different age groups to transmission, and the dashed lines show the 95% posterior credible intervals for ℛ_*e*_(*t*). The vertical dotted line indicating the introduction of the first lockdown (LD 1) coincides with the start of CP1.

Figure 3**B** shows that if both educational levels had remained open, the peak in hospital bed occupancy would have been much higher than in the baseline scenario, and the hospitalization wave would not have subsided. Keeping only elementary schools or only secondary schools open would have resulted in peaks of similar height, both exceeding that of the baseline scenario. However, the wave would have been more prolonged when secondary schools remained open, indicating a more sustained burden on the healthcare system compared to the scenario with open elementary schools.

To further investigate the model dynamics in CP1, we computed the time-varying reproduction number, ℛ_*e*_(*t*), and cumulative elasticities quantifying the contributions of children, adolescents, and adults to transmission (Figure 3**C**-**F**). Following the implementation of the first lockdown in the baseline scenario, ℛ_*e*_(*t*) declined from 2.5 (95% CrI 2.2–2.7) and remained below or close to 1, consistent with the curbed wave of hospitalizations shown in Figure 3**B**. With schools closed, adults were the primary contributors to transmission, accounting for at least 80% of the cumulative elasticities (Figure 3**C**). In contrast, if schools had remained open, ℛ_*e*_(*t*) would have increased despite the lockdown introduction, reaching 2.5 (95% CrI 2.1–3.0) when both school types were open, 2.1 (95% CrI 1.7–2.6) when only secondary schools were open, and 1.8 (95% CrI 1.3–2.3) when only elementary schools were open. These findings align with the respectively larger hospitalization waves in Figure 3**B**. When secondary schools remained open, either alone or together with elementary schools, the largest contribution to transmission shifts from adults to adolescents (Figure 3**D** and **F**). In the scenario of only opening elementary schools (Figure 3**E**), children play a large role in transmission, although their contribution would be smaller than that of adolescents in the scenario with open secondary schools.

### Second counterfactual period: CP2

In CP2, we explored the opposite scenario: the impact of closing schools when they were, in fact, open (Figure 4). We focused our analysis on the period following summer school holidays to evaluate whether school closures could have prevented the autumn wave of hospitalizations in 2020. As shown in Figures 4**A**-**B**, closing both elementary and secondary schools would have prevented a surge in hospitalizations, reducing total cumulative hospitalizations relative to the baseline by 87% (95% CrI 79%–90%). A comparable but smaller reduction would have been achieved if only secondary schools had been closed (75% decrease, 95% CrI 58%–82%), suggesting that interventions targeting this type of schools are quite effective in curbing transmission. In contrast, closing only elementary schools would have led to a more modest reduction in hospitalizations (51%, 95% CrI 42%–58%).

**Figure 4.**
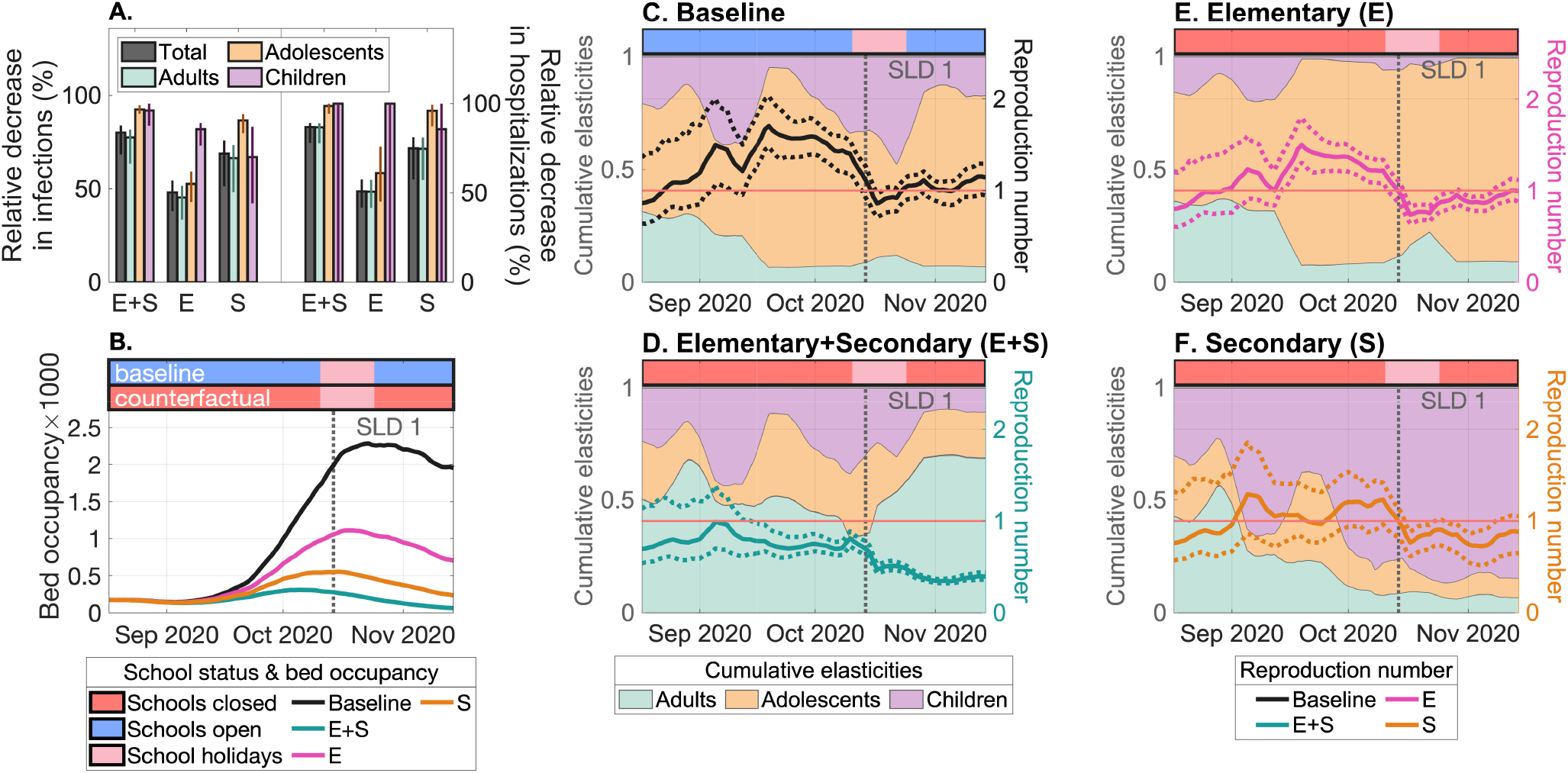
Retrospective evaluation of closing elementary and secondary schools in CP2. The colored horizontal bars in (**B**)-(**F**) indicate the status of school operations in the baseline and counterfactual scenarios. **(A)** Relative decrease in cumulative infections and hospitalizations in the counterfactual scenarios compared to the baseline, by age group and for the total population. The bars represent median values, and the whiskers indicate the 95% posterior credible intervals. **(B)** Projected median hospital bed occupancy over time for the baseline and counterfactual scenarios. (**C**)-(**F**) Mean cumulative elasticities by age group (left y-axis) and median effective reproduction number (right y-axis) for **(C)** the baseline and three counterfactual scenarios: **(D)** full school closure, **(E)** closure of elementary schools only, and **(F)** closure of secondary schools only. In (**C**)-(**F**) the shaded areas represent the contributions of different age groups to transmission, and the dashed lines show the 95% posterior credible intervals for ℛ_*e*_(*t*). The vertical dotted line indicates the introduction of the first semi-lockdown (SLD 1).

In the baseline scenario with open schools in CP2 (Figure 4**C**), ℛ_*e*_(*t*) begins to rise above 1 in September 2020 and only drops below 1 following the introduction of the first semi-lockdown. In this scenario, adolescents were the primary drivers of transmission, followed by children and, to a much smaller extent, adults. The counterfactual closure of both elementary and secondary schools — representing the intervention with the largest impact on total hospitalizations — reduces ℛ_*e*_(*t*) and maintains it below one throughout CP1, in line with the absence of a substan-tial hospitalization wave (Figure 4**D**). In this scenario, the relative contribution of adults to transmission markedly increases, particularly after the semi-lockdown is implemented, as transmission among children and adolescents is suppressed. By contrast, closing only elementary schools (Figure 4**E**) or only secondary schools (Figure 4**F**) fails to bring ℛ_*e*_(*t*) below 1, resulting in hospitalization waves shown in Figure 4**B**. In these scenarios, transmission remains predominantly driven by adolescents when only elementary schools are closed and by children when only secondary schools are closed, underscoring the important roles of these groups in sustaining transmission.

### Third counterfactual period: CP3

The analysis for CP3, spanning 4 January 2021 to 7 February 2021, evaluates the impact of fully reopening schools after the Christmas holidays in 2020. As a reminder, during this period, secondary schools operated at 25% capacity and elementary schools were closed. Fully opening both educational levels would have led to a 129% (95% CrI 93%– 168%) increase in total hospitalizations, triggering a new large wave (Figure 5**A** and5**B**). Opening only secondary or only elementary schools would have resulted in a similar but smaller wave, with total hospitalizations increasing by 48% (95% CrI 36%–67%) and 51% (95% CrI 28%–81%), respectively. Unlike in CP1 and CP2, where secondary schools had a larger impact on transmission than elementary schools, the effects of opening both elementary and secondary schools in CP3 were more comparable. This is expected, as adolescents were already playing a role in transmission under the baseline scenario due to the partial reopening of secondary schools.

**Figure 5.**
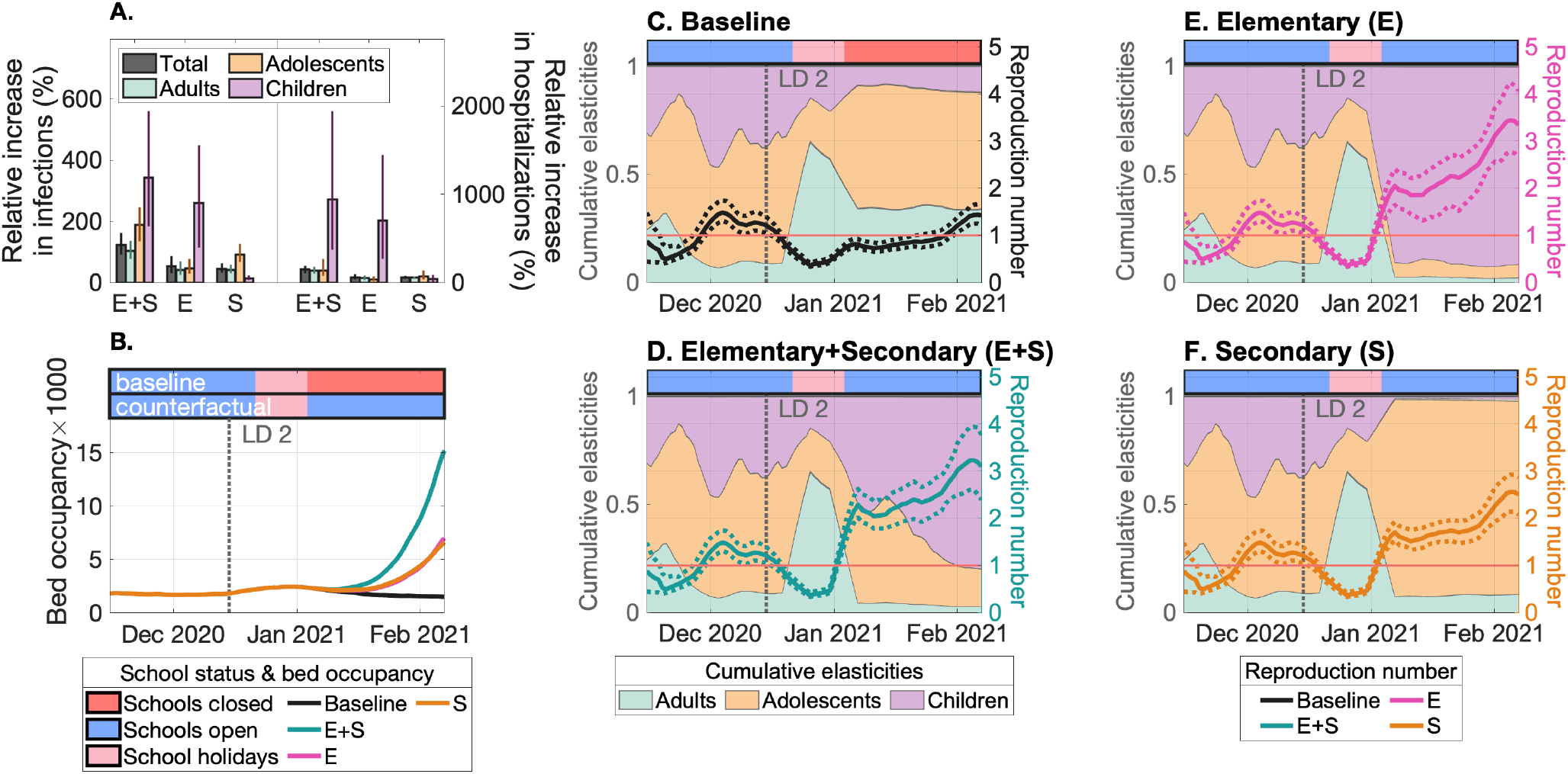
Retrospective evaluation of opening elementary and secondary schools in CP3. The colored horizontal bars in (**B**)-(**F**) indicate the status of school operations in the baseline and counterfactual scenarios. **(A)** Relative increase in cumulative infections and hospitalizations in the counterfactual scenarios compared to the baseline, by age group and for the total population. The bars represent median values, and the whiskers indicate the 95% posterior credible intervals. **(B)** Projected median hospital bed occupancy over time for the baseline and counterfactual scenarios. (**C**)-(**F**) Mean cumulative elasticities by age group (left y-axis) and median effective reproduction number (right y-axis) for **(C)** the baseline and three counterfactual scenarios: **(D)** full school opening, **(E)** opening of elementary schools only, and **(F)** opening of secondary schools only. In (**C**)-(**F**) the shaded areas represent the contributions of different age groups to transmission, and the dashed lines show the 95% posterior credible intervals for ℛ_*e*_(*t*). The vertical dotted line indicates the introduction of the second lockdown (LD 2).

In the baseline scenario for CP3, adolescents and children were the main contributors to transmission until Christmas holidays (Figure 5**C**). After the start of the second lockdown, ℛ_*e*_(*t*) dropped below 1, accompanied by an abrupt increase in adult cumulative elasticities, owing to school closures during the winter break. After partial reopening of secondary schools and continued closure of elementary schools, ℛ_*e*_(*t*) stayed below 1 until 25 January 2021, with adolescents consistently displaying the highest cumulative elasticities among age groups. The gradual rise in ℛ_*e*_(*t*) toward the end of CP3 reflects the spread of the Alpha variant of concern. In the counterfactual scenarios where elementary schools were reopened (E+S and E), peak ℛ_*e*_(*t*) values would have exceeded 3, with children clearly driving transmission during this period (Figure 5**D** and 5**E**). These resulting high ℛ_*e*_(*t*) values align with high children contacts and a rapidly decreasing proportion of susceptible children (Supplementary Figures S4 and S16). Reopening only secondary schools would have led to adolescents becoming the primary contributors to transmission (Figure 5**F**).

### Fourth counterfactual period: CP4

Similar to CP2, CP4, which includes the second semi-lockdown, examines the effects of school closures during November-December 2021, when schools were actually open until they closed for the winter holidays. Closing both educational levels would have resulted in the largest reduction in total hospitalizations, i.e., 60% (95% CrI 58%–63%) (Figure 6**A**). A similar observation was done in CP2, however unlike in CP2, the impact of widespread school closures on hospitalizations is smaller because of the uptake of COVID-19 vaccination in the population (Supplementary Figure S14). Also unlike in CP2, closing elementary schools in CP4 would have been more impactful than closing secondary schools (38% vs 31% reduction in total hospitalizations). This difference is also evident in Figure 6**B**, which shows higher hospital bed occupancy when only secondary schools are closed compared to when only elementary schools are closed. This result is primarily explained by the start of COVID-19 vaccination among adolescents in July 2021, whereas no children had been vaccinated by that time. In addition, a higher proportion of adolescents than children had previously experienced primary infection, providing them with partial immunity (Supplementary Figure S15). In contrast, unvaccinated and infection-naïve children remained fully susceptible and fully infectious upon infection.

**Figure 6.**
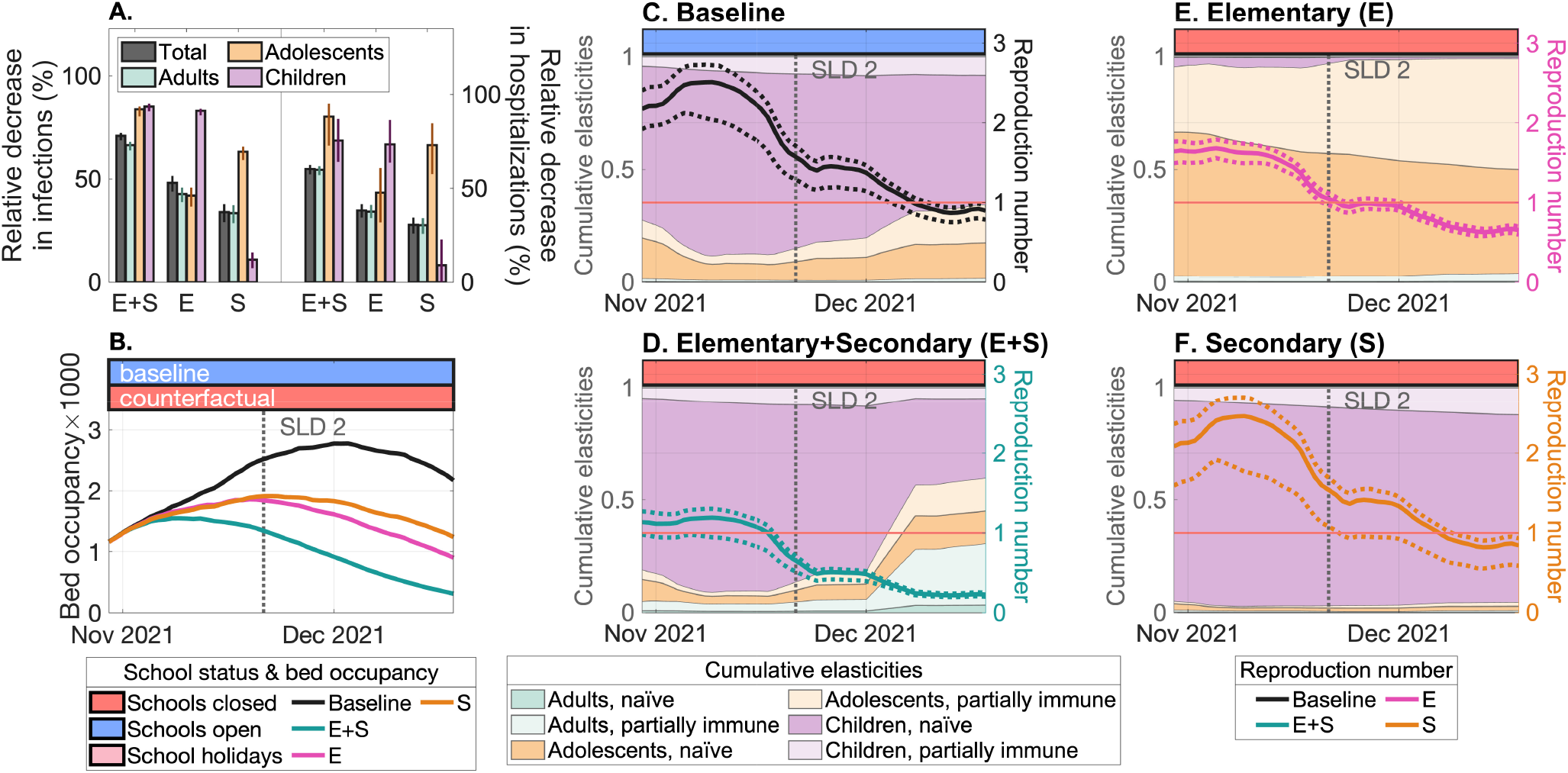
Retrospective evaluation of closing elementary and secondary schools in CP4. The colored horizontal bars in (**B**)-(**F**) indicate the status of school operations in the baseline and counterfactual scenarios. **(A)** Relative decrease in cumulative infections and hospitalizations in the counterfactual scenarios compared to the baseline, by age group and for the total population. The bars represent median values, and the whiskers indicate the 95% posterior credible intervals. **(B)** Projected median hospital bed occupancy over time for the baseline and counterfactual scenarios. (**C**)-(**F**) Mean cumulative elasticities by age group (left y-axis) and median effective reproduction number (right y-axis) for **(C)** the baseline and three counterfactual scenarios: **(D)** full school closure, **(E)** closure of elementary schools only, and **(F)** closure of secondary schools only. In (**C**)-(**F**) the shaded areas represent the contributions of different age groups to transmission, and the dashed lines show the 95% posterior credible intervals for ℛ_*e*_(*t*). The vertical dotted line indicates the introduction of the second semi-lockdown (SLD 2).

We confirm this in Figure 6**C**, where in the baseline scenario with fully opened schools, naïve children dominated transmission throughout the period, accounting for over 68% of the cumulative elasticities. CP4 marked the first time that adults had a minimal role in transmission, while contributions from partially immune adolescents and children began to emerge. In the counterfactual scenario with all schools closed (Figure 6**D**), ℛ_*e*_(*t*) declined relative to the baseline scenario, yet naïve children remained the primary contributors to transmission. Closing only secondary schools (Figure 6**F**) similarly resulted in a children-driven pandemic, with ℛ_*e*_(*t*) values comparable to the baseline scenario. In contrast to these three scenarios (Baseline, E+S, and E), closing only elementary schools (Figure 6**E**), shifted the dominant source of transmission to adolescents, both naïve and partially immune.

## Discussion

To our knowledge, this is the first study to systematically evaluate how alternative school closure and reopening strategies for elementary and secondary schools could have influenced the course of the COVID-19 pandemic during 2020–2021, and one of the most detailed national-level assessments of school closures to date. During the first wave in spring 2020 (CP1), our counterfactual analysis suggests that keeping all schools open with pre-pandemic contact patterns would have led to widespread transmission and a major surge in hospitalizations. This underscores the critical role of school closures in the early phase of the pandemic, a period with limited alternative interventions, in averting an unmanageable burden on the healthcare system. In the period following the summer holidays of 2020 (CP2), our findings indicate that maintaining schools closed for in-person education could have prevented the resurgence of hospitalizations observed in the autumn of 2020. This highlights a missed opportunity for mitigating transmission through more cautious reopening of schools. In CP3, the decision to keep schools closed after the Christmas break in late 2020 appears to have been effective in limiting a hospitalization wave associated with the Alpha variant. Reopening all schools at that time would likely have triggered a major increase in transmission and hospital burden. Finally, schools remained open during CP4, the end of 2021, despite rising hospitalizations. Our counterfactual scenarios suggest that closing schools during this period could have contributed meaningfully to reducing the magnitude of the late 2021 wave. Overall, these findings indicate that school interventions, while highly context-dependent, had the potential to mitigate transmission and hospitalization burden across multiple phases of the pandemic, and that targeted use of school closures might have altered the trajectory of the pandemic in the Netherlands.

Comparing our findings with those of previous studies is challenging, as most evaluations of school interventions have relied on forward projections rather than counterfactual analyses (e.g., [28, 64–66]). Despite variation in study design, scope, and setting, the broader body of evidence suggests that school closures tend to reduce SARS-CoV-2 transmission, which aligns with our findings [28, 29, 31, 36, 38]. A more direct comparison is possible with Ragonnet et al. [38], who conducted a counterfactual analysis to evaluate the impact of keeping all schools open throughout 2020–2022. Their findings for the first wave are broadly consistent with ours, particularly regarding the peak in hospital bed occupancy. However, an agent-based modeling study by Dekker et al. [37] found that school closures would have had a small impact on the first wave. The basis for this discrepancy is not entirely clear, but it may reflect differences in the timing of school closures and their assumed effects on contact patterns across age groups in the respective models.

In contrast to most previous modeling studies [28, 36, 38, 67, 68], our analysis distinguishes between elementary and secondary schools, allowing us to assess their individual and combined contributions to transmission dynamics. Across all counterfactual periods, the simultaneous closure or reopening of both educational levels consistently had the largest impact on transmission. However, due to the nonlinear nature of the model, the combined effect was not simply the sum of the individual effects. In other words, simultaneous closing (or opening) of both school types did not equal the sum of closing (or opening) each educational level independently. During the earlier phases of the pandemic (CP1 and CP2), interventions targeting secondary schools had a stronger impact than those targeting elementary schools, producing effects comparable to an intervention targeting both school types. This likely reflects a larger number of contacts in secondary school settings and a higher susceptibility of adolescents relative to children. In later phases (CP3 and CP4), however, this pattern reversed: closures and openings of elementary schools had a larger effect than those of secondary schools. This shift can be explained by differences in operational status (i.e., secondary schools were already partially open in CP3) and population immunity, where fewer children than adolescents had acquired partial protection through prior infection, and very few had been vaccinated by the end of 2021 (CP4). These findings demonstrate that the impact of school interventions is not fixed but depends on several factors, including school type, age-specific population immunity, contact patterns, and the timing of policies. As such, decisions around school interventions should be carefully tailored to the specific epidemiological context, particularly in pandemics where these factors shift over time.

While our findings offer insights into SARS-CoV-2 transmission, they should not be directly generalized to other respiratory pathogens. The role of schools in disease spread is contingent on age-specific epidemiological parameters, such as susceptibility to infection, infectivity, and disease severity. In the case of the 2009 influenza pandemic, for instance, children were thought to contribute substantially to transmission, partly due to their high contact rates and increased susceptibility to the virus [69,70]. Although our modeling framework is adaptable and could be applied to evaluate school openings and closures for other pathogens with pandemic potential, the specific findings presented here are tailored to the epidemiology of SARS-CoV-2. Future policy decisions should therefore be informed by pathogen-specific data and context, rather than assuming uniform effects of school strategies across respiratory diseases.

Our study goes beyond projecting the impact of school interventions on infections, hospitalizations, and the reproduction number, as commonly done in previous work [33, 71, 72]. We used elasticity analysis, as developed in [73], and applied it to the time-varying effective reproduction number to quantify the contributions of different age groups and immunity classes to transmission across all periods. This analysis is essential for identifying the age groups that drive transmission and for interpreting the results of counterfactual scenarios. During the first wave in spring 2020, adults were identified as the primary drivers. In subsequent periods, however, children and adolescents played an increasingly important role in sustaining SARS-CoV-2 spread. Adolescents contributed more prominently than children in the autumn of 2020 and early 2021, while children emerged as the main drivers of transmission in the final counterfactual period. By the end of 2021, our elasticity analyses also revealed that population immunity had become an important factor in shaping transmission dynamics, with an increasing share of transmission attributed to partially immune children and adolescents. This finding aligns with previous studies for the United Kingdom [74] and the Netherlands [53], which showed a dramatic increase in reinfections in late 2021. Our results from elasticity analyses are broadly consistent with previous studies for Belgium [50, 52] and Portugal [36, 51], which applied the same methodology, albeit at a coarser or shorter temporal scale. Common findings include adult-driven transmission during the first wave and high cumulative elasticities among children during late 2021, particularly in Belgium [52], with the overall contribution of school-aged populations to transmission varying based on country-specific context.

Some simplifications were made in the model structure to tailor it for our research question. We did not include seasonality in transmission, although it can be readily incorporated into the model. Our previous work [53] found that a model without seasonality provided a better fit to the data, suggesting that non-pharmaceutical interventions were the dominant drivers of transmission during the study period. Individuals who had acquired partial immunity, either through prior infection or vaccination, were grouped because the available vaccination data did not distinguish between different pathways to partial immunity (e.g., two doses of mRNA vaccines, one vaccine dose plus prior infection, or a single Janssen dose). The compartmental nature of the model makes it less suitable for explicitly capturing granular school-based interventions, such as hybrid learning, mask-wearing, smaller class sizes, and regular testing, which were evaluated in other modeling studies specifically designed for such interventions [43–45, 68, 72]. Finally, to balance data limitations and model complexity, we stratified the population into three broad age groups—children (0–9 years), adolescents (10–19 years), and adults (20+ years). This decision was informed by constraints on the number of estimated age-specific parameters, the available contact matrices stratified in 10-year age bands, and privacy restrictions associated with hospitalization data. While this approach enabled robust model fitting and interpretation, it did not allow for a fully granular attribution of transmission to elementary (up to 12 years) versus secondary (13–19 years) school-aged populations.

Looking ahead, our model could be extended to explore which levels of non-school contacts might have allowed schools to remain open without overwhelming the healthcare system. Such analyses could help identify the timing and magnitude of non-school interventions necessary to offset transmission risks in schools and to prevent their closures during different phases of the pandemic. They would support preparedness planning by providing better guidance on how to balance school operations with other control measures. Another important direction for future work is to integrate societal consequences of school closures into modeling studies. School closures not only reduce transmission but also have adverse effects on learning, mental health, and well-being of children [24, 75–79]. Incorporating these non-epidemiological impacts would inform more balanced decision-making regarding school closure policies in future pandemics.

## Conclusions

In conclusion, by combining counterfactual analysis with time-varying elasticity measures, we showed that the effectiveness of school interventions was highly context-dependent, shaped by factors such as school type, contact patterns, policy timing, and population immunity. The study also highlighted shifting contributions of different age groups to transmission over time, i.e., adults were the main drivers early in the pandemic, followed by adolescents in late 2020 and early 2021, and children in late 2021. Reflecting these dynamics, the relative importance of school types also changed with time. Secondary schools had a greater impact in 2020, while elementary schools played a larger role in 2021. Our findings support a more targeted and adaptive approach to school closures and openings in future pandemics.

## Supplementary information

The Supplementary Material provides additional figures, tables, and further details of this study.

## Acknowledgements

The authors gratefully acknowledge funding from the Fundącão para a Ciência e a Tecnologia, I.P., through national funds, under the project 2022.01448.PTDC, DOI 10.54499/2022.01448.PTDC. This work was also supported by UID/04046/2025, Instituto de Biosistemas & Ciências Integrativas Centre grant from FCT, Portugal. Benedetta Canfora was supported by the Swaantje Mondt travel fund from the Center for Complex Systems Studies at Utrecht University. Ganna Rozhnova was supported by the VERDI project (101045989), funded by the European Union. Views and opinions expressed in this article are however those of the author(s) only and do not necessarily reflect those of the European Union or the Health and Digital Executive Agency. Neither the European Union nor the granting authority can be held responsible for them. Otilia Boldea was supported by Tilburg University Fund. We thank Prof. Marc Bonten (University Medical Center Utrecht), Dr Susan van den Hof and Dr Jantien Backer (National Institute for Public Health and the Environment), Prof. Dennis Huisman (Erasmus University Rotterdam), and Jan Banninga (Nederlandse Spoorwegen) for their substantial help in obtaining the data for this study. We also thank the members of the Infectious Disease Modeling Group (University Medical Center Utrecht), Dr Sen Pei and Prof. Jeffrey Shaman (Columbia University) for useful discussions on data availability and modeling.

## Author contributions

Conceptualization: G.R. and O.B.; Methodology: G.R. and O.B.; Software: B.C., R.A.E., and O.B.; Validation: O.B. and A.N.; Formal Analysis: B.C., R.A.E., G.R., and O.B.; Investigation: B.C., R.A.E., G.R., and O.B.; Resources: G.R. and A.N.; Data Curation: O.B.; Writing–Original Draft: B.C., R.A.E., and G.R.; Writing–Review & Editing: All authors; Visualization: R.A.E.; Supervision: G.R., O.B., and A.N.; Project Administration: G.R. and A.N.; Funding Acquisition: G.R.

## Data availability

All datasets analyzed and generated in this study will be available upon publication on https://github.com/becanfora/COVID-NL-Counterfactual. The contents of the GitHub repository are available upon request for peer review.

## Code availability

The codes reproducing the results of this study will be available upon publication on https://github.com/becanfora/COVID-NL-Counterfactual. The contents of the GitHub repository are available upon request for peer review.

## Supplementary Material

### S1 Model equations and parameters

The age-structured metapopulation model used in this study, as discussed in Section and visualized in Figure 1, is based on the work of [1]. The system of equations is given by Equation S1.

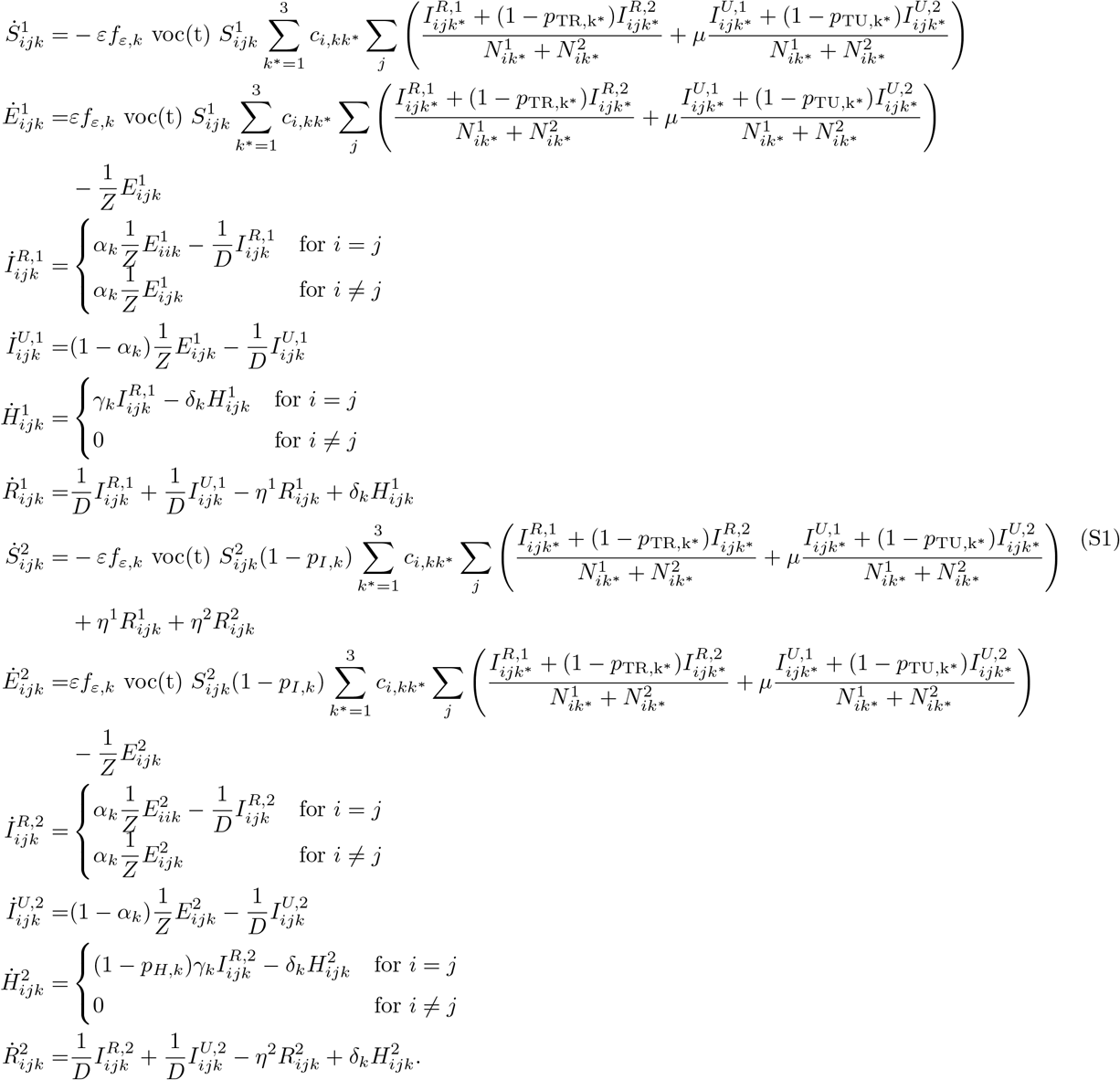

The compartmental model distinguishes individuals who are susceptible (*S*), exposed (*E*), reported infected (*I*^*R*^), unreported infected (*I*^*U*^), hospitalized (*H*), and recovered (*R*). The total population is denoted by *N*. Since the model assumes constant demography, the sum of all population compartments equals *N*. The immunity status of each population is described by superscript *a* = 1, 2. *a* = 1 denotes naïve transmission dynamics, while *a* = 2 corresponds to partial immunity resulting from prior infection or vaccination. The structural components of the model are defined by subscripts, e.g., *i* = 1, …, 12 for destination region (twelve Dutch provinces), *j* = 1, …, 12 for origin region (twelve Dutch provinces), and *k* = 1, 2, 3 for age group (adults, adolescents, and children).

### S2 Contact patterns

**Table S1.** Timeline of contact data surveys. Shown are the minimum, maximum, mean, and median times for each of the eleven surveys.

| Survey | Minimum time | Maximum time | Mean time | Median time |
| --- | --- | --- | --- | --- |
| Survey 1 | 02/2016 | 10/2017 |  |  |
| Survey 2 | 30/03/2020 | 13/05/2020 | 21/04/2020 | 02/04/2020 |
| Survey 3 | 08/06/2020 | 10/08/2020 | 10/07/2020 | 13/06/2020 |
| Survey 4 | 21/09/2020 | 17/12/2020 | 04/11/2020 | 27/09/2020 |
| Survey 5 | 10/02/2021 | 02/10/2021 | 22/03/2021 | 16/02/2021 |
| Survey 6 | 14/06/2021 | 19/08/2021 | 17/07/2021 | 22/06/2021 |
| Survey 7 | 31/10/2021 | 20/01/2021 | 10/12/2021 | 10/11/2021 |
| Survey 8 | 12/03/2022 | 24/05/2022 | 17/04/2022 | 22/03/2022 |
| Survey 9 | 13/06/2022 | 24/10/2022 | 18/08/2022 | 20/06/2022 |
| Survey 10 | 29/10/2022 | 09/01/2023 | 04/12/2022 | 08/11/2022 |
| Survey 11 | 16/04/2023 | 01/06/2023 | 09/05/2023 | 24/07/2023 |

Shown in Figure S1 are the timelines for each counterfactual period (CP1–CP4), indicating the dates at which changes in contact patterns occurred, the specific levels at which these changes took place, and the corresponding school operational status (i.e., open, closed, holidays, or partially open). Each period is associated with a specific contact matrix that gives the total number of contacts. From this total, a fraction *q* is estimated to calculate the number of school contacts, expressed as a fraction of pre-pandemic school contacts. The total number of contacts is then the sum of school and non-school contacts. The value of *q* varies across surveys and periods, as described in the Methods section of the main text.

All contact matrices (Figure S2) are further scaled by a school opening index (Figure S3), which reflects the level of school opening or closure in each province. Specifically, when schools are closed, the school contact matrices are multiplied by 0, effectively nullifying school contacts. When schools are fully open, the matrices are multiplied by 1. For partial openings, a proportional factor is applied based on school capacity. This index allows us to model different scenarios, as it is the primary varying parameter across simulations. To show how contacts vary across periods in both the baseline and counterfactual scenarios, we plot the age-specific average contacts for South Holland in Figure S4.

**Figure S1.**
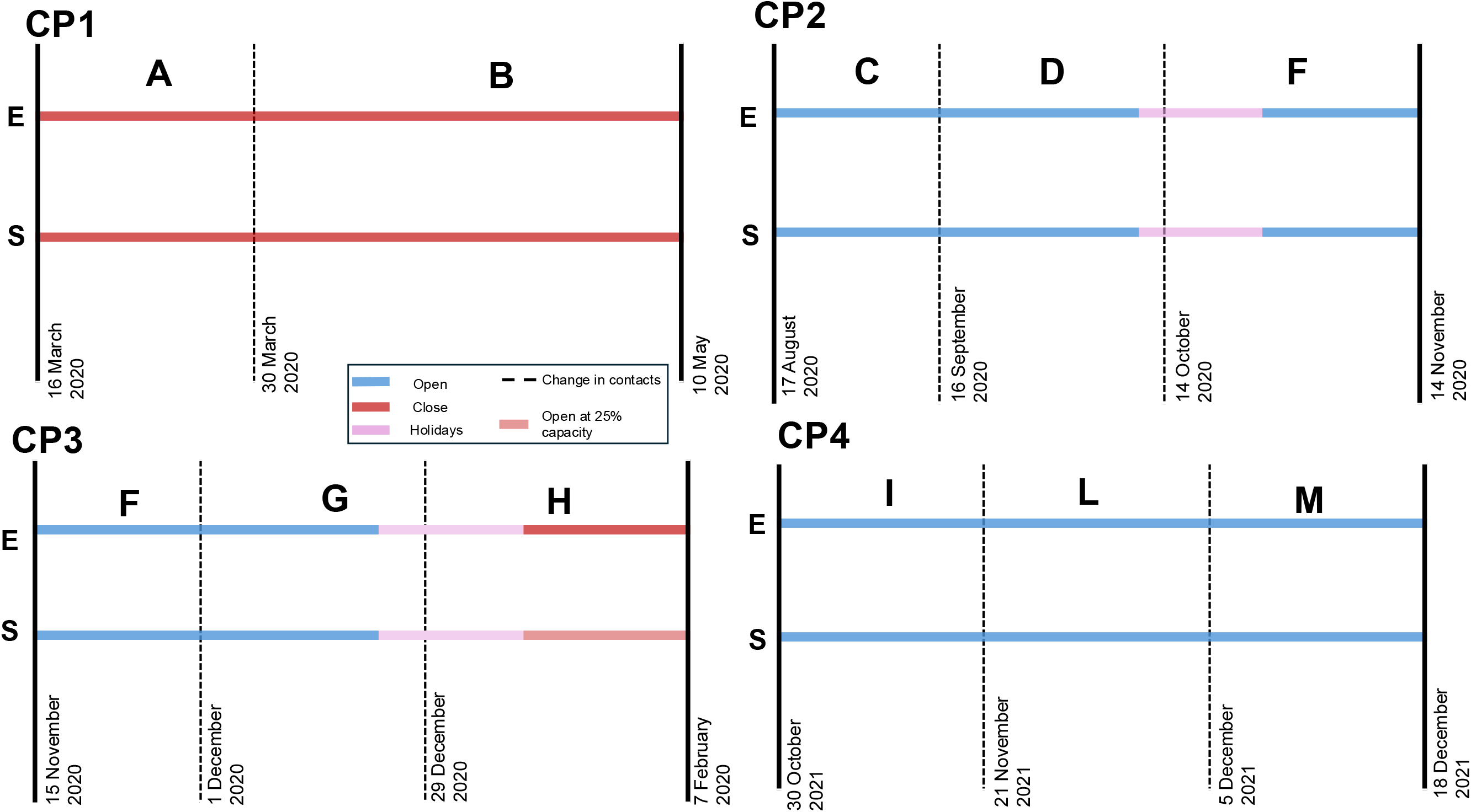
Timeline of elementary (E) and secondary (S) school operational status across all periods (CP1–CP4). The red lines indicate school closures, blue lines represent school openings, and pink segments denote holiday periods. Light red indicates partial reopening at 25% capacity. Dashed vertical lines mark changes in contact matrices.

**Figure S2.**
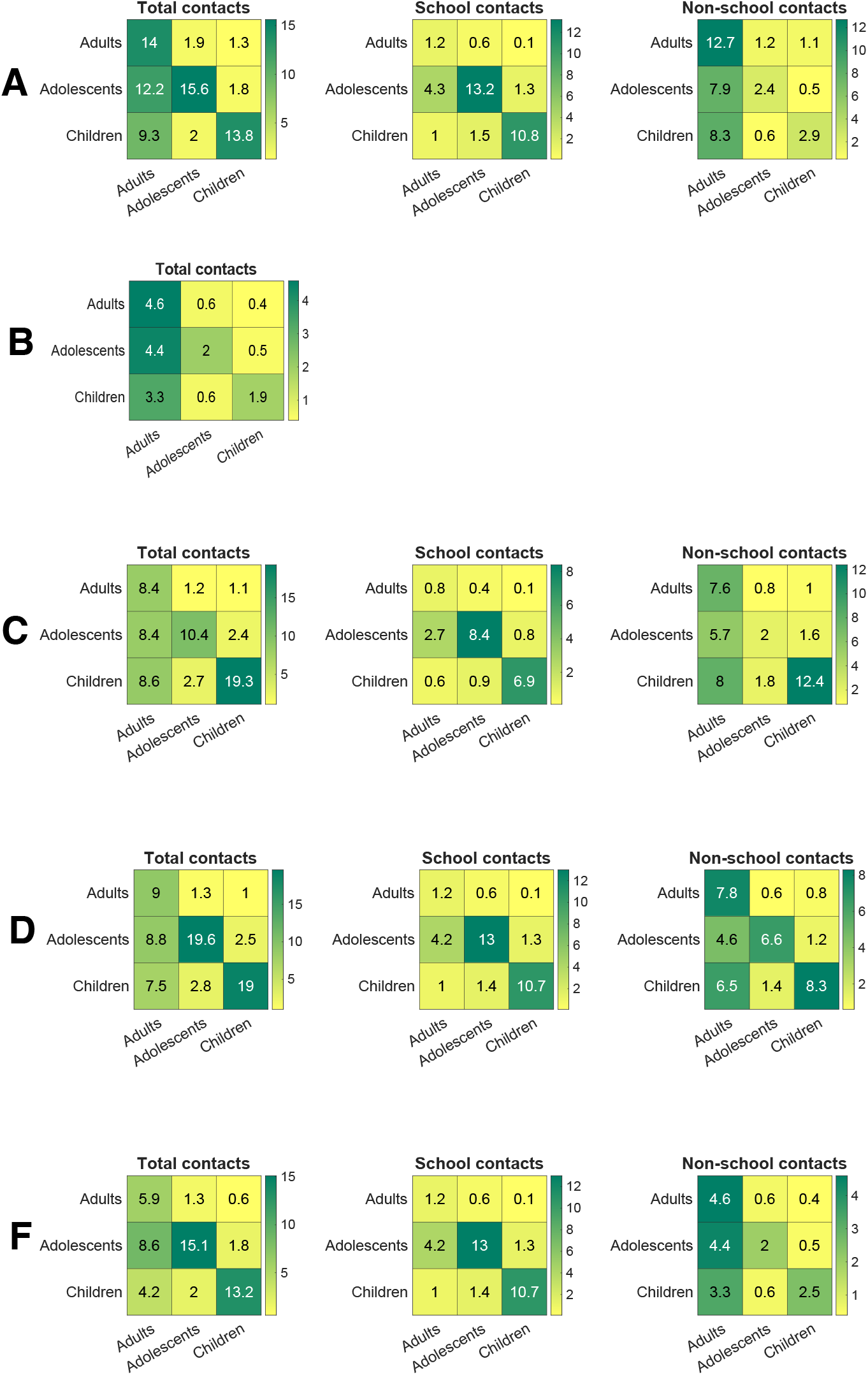
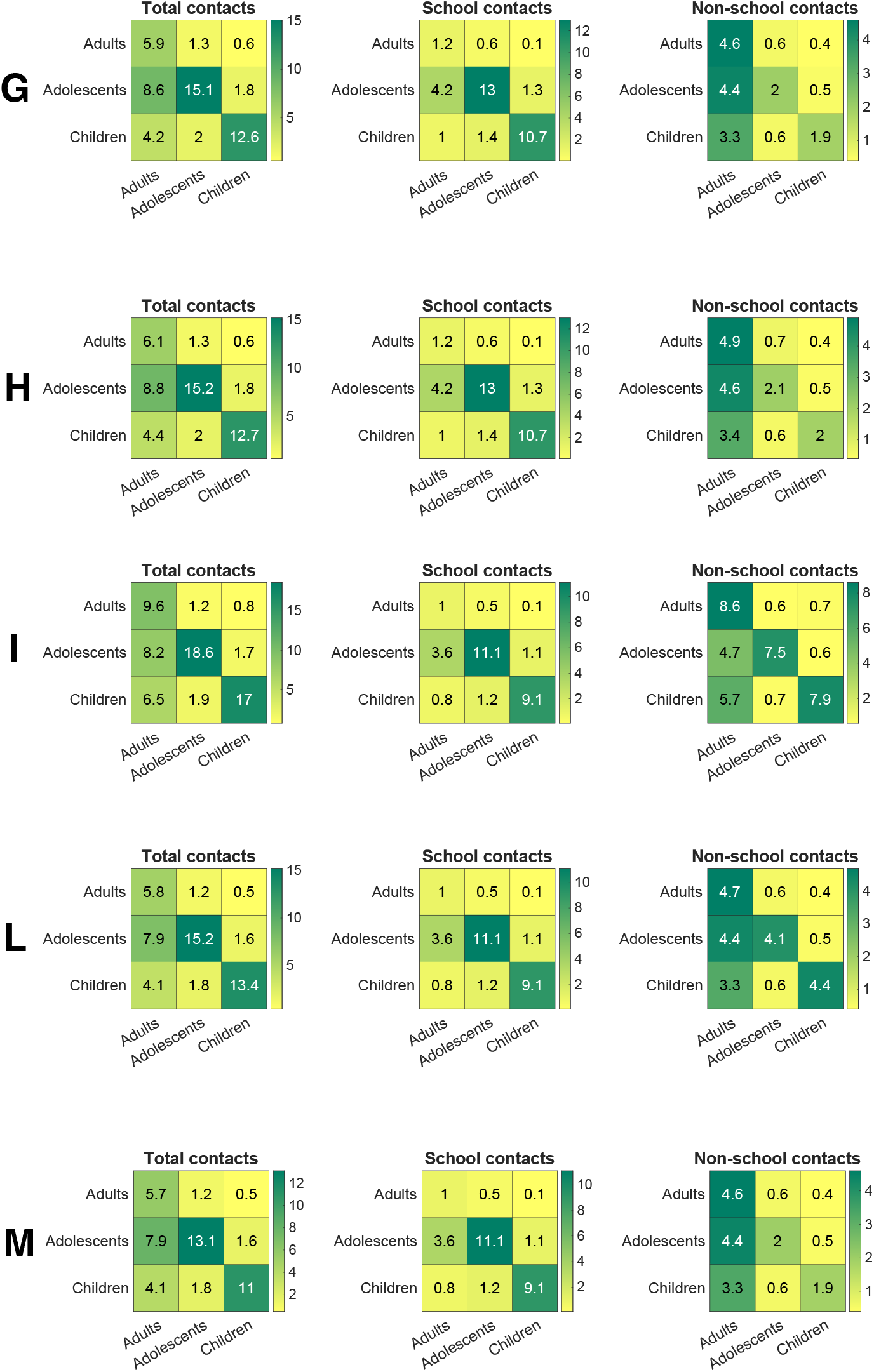
Contact matrices used in the analysis. Each panel shows total contacts (left), school contacts (middle), and non-school contacts (right), stratified by age group (adults, adolescents, children). **A** Pre-pandemic contact patterns from Survey 1. School contacts derived directly from data. **B** Total contacts during Survey 2, corresponding to the first lockdown period. **C** Contacts from Survey 3; school contacts computed using a fraction *q* = 0.636 of the pre-pandemic school contact matrix. **D** Contacts from Survey 4; school contacts computed with *q* = 0.9890. **F** Same school contact matrix as in **D**, but non-school contacts reduced by a factor of 0.3 (value determined via grid search to minimize distance between simulated and actual infections and hospitalizations).Contact matrices used in the analysis. Each panel shows total contacts (left), school contacts (middle), and non-school contacts (right), stratified by age group (adults, adolescents, children). **G** Contacts from Survey 2 (lockdown), combined with school contacts from **D. H** Same as **G**, but with non-school contacts increased by a factor of 1.05. **I** Contacts from Survey 7; school contacts computed with *q* = 0.8390. **L** Same school contacts as in **I**, but with non-school contacts reduced by a factor of 0.55. **M** Contacts from Survey 2, combined with school contacts from Survey 7.

**Figure S3.**
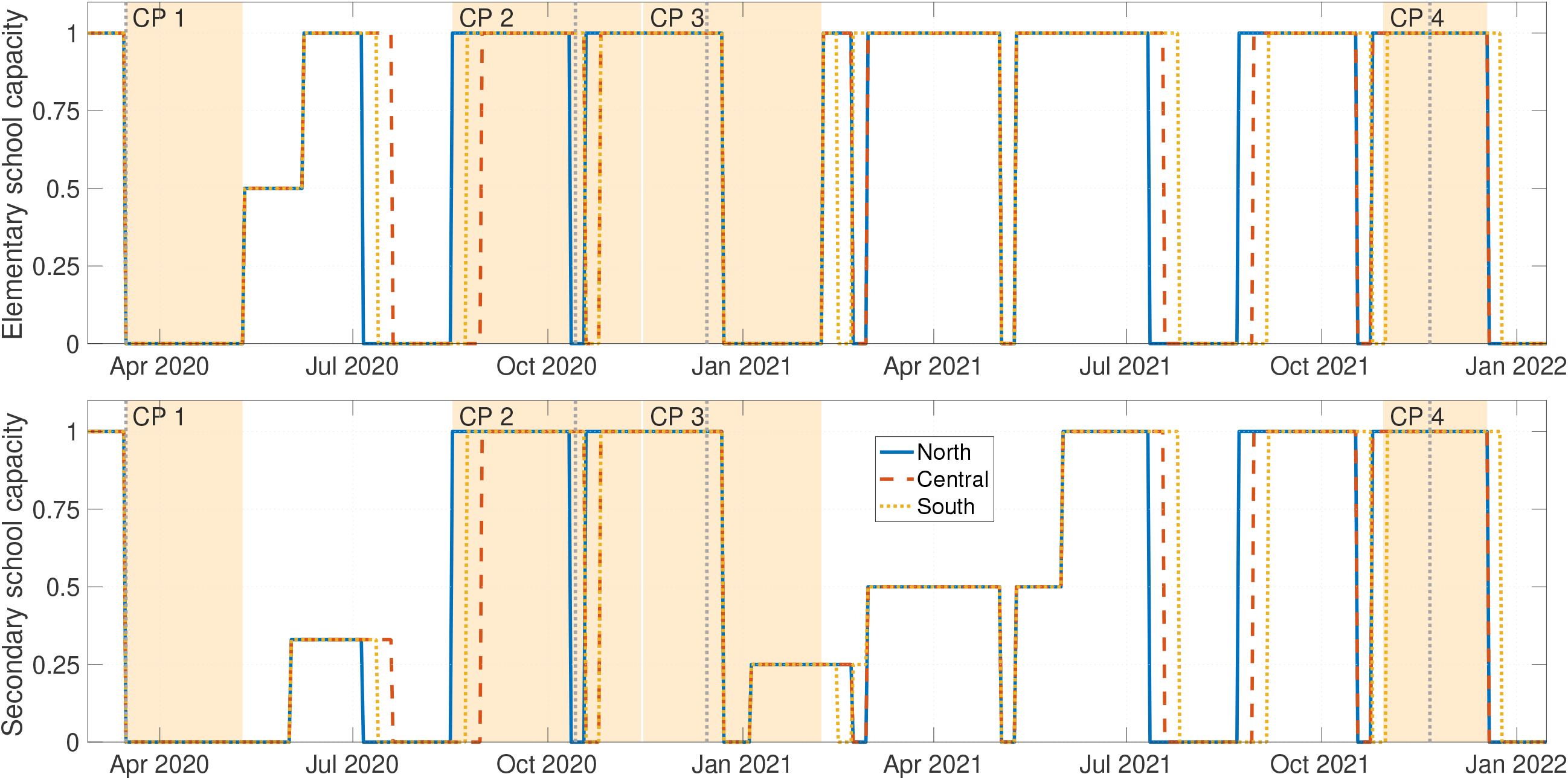
Elementary and secondary school capacity. School indices for (top) elementary and (bottom) secondary school openings in the North, Central, and South provinces in the Netherlands. The northern regions (blue solid) include Groningen, Friesland, Drenthe, Overijssel, Flevoland, and North Holland. The central regions (red dashed) include Gelderland, Utrecht, and South Holland. The southern regions (yellow dotted) include Zeeland, North Brabant, and Limburg. The yellow-shaded regions mark the periods used for counterfactual analyses. The vertical dotted line indicates the introduction of lockdowns and semi-lockdowns corresponding to the counterfactual periods. CP: counterfactual period.

**Figure S4.**
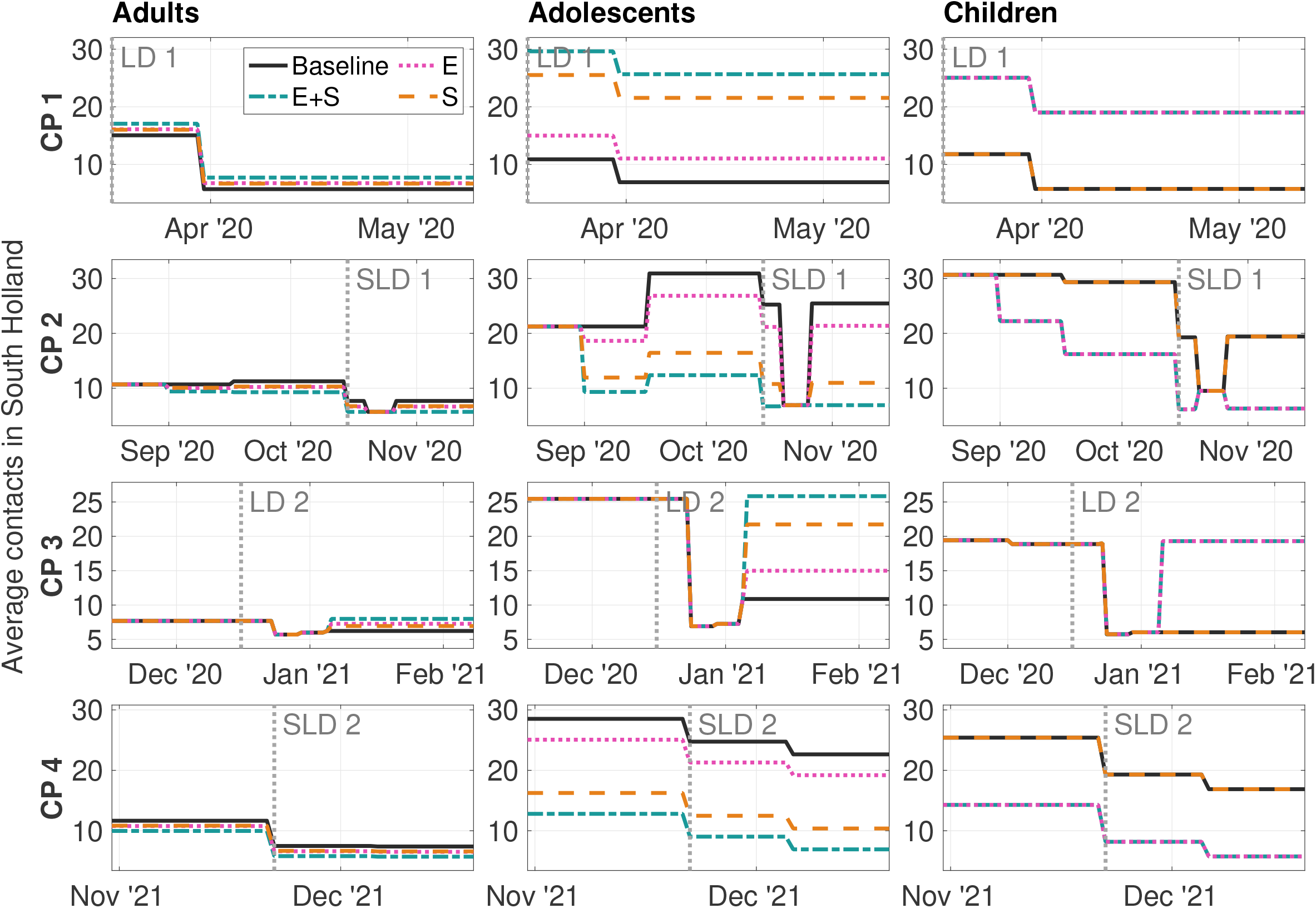
Contact rates over time in South Holland. Average number of daily contacts in adults, adolescents, and children for all periods considered for the baseline and counterfactual scenarios. Lines correspond to the baseline (black solid), counterfactual scenario of both elementary and secondary schools (E+S, blue green dashed-dotted), counterfactual scenario of elementary schools only (E, pink dotted), and counterfactual scenario of secondary schools only (S, orange dashed). The vertical dotted line indicates the introduction of lockdowns and semi-lockdowns corresponding to the counterfactual periods. CP: counterfactual period. LD: lockdown. SLD: semi-lockdown.

### S3 Parameter posterior distributions

Model parameters are detailed in Table S2, which specifies how they are defined for each period. A parameter not explicitly shown in the table is the contact rate, 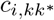 (*t*), which quantifies the rate of contact between an individual in age group *k*^*^ and another individual in age group *k* within region *i*. Furthermore, the model incorporates a logistic function for variants of concern, voc(t), that models an increase in the probability of transmission due to new variants. The construction of this function is thoroughly described in [1].

The parameter posterior distributions at the end of the period compared to their prior ranges are shown in Figures S5-S8. Values of parameters that were not estimated are given in Table S2. In particular, *µ, D*, and *Z* were calibrated based on the average posteriors over time in [1] (Supplementary Materials, Figure A2). The calibration of *θ*_1_ relies on the average percentage increase in Google mobility data relative to train mobility data for adults, while for adolescents and children we assumed much lower commuting frequencies than for adults. Fixed parameters were selected through grid searches, ensuring that the baseline counterfactual simulations (using the estimated model parameters) closely matched the observed data. Specifically, parameters were chosen to minimize the median distance between simulated infections and hospitalizations in the baseline scenario and the corresponding empirical data.

**Table S2.**
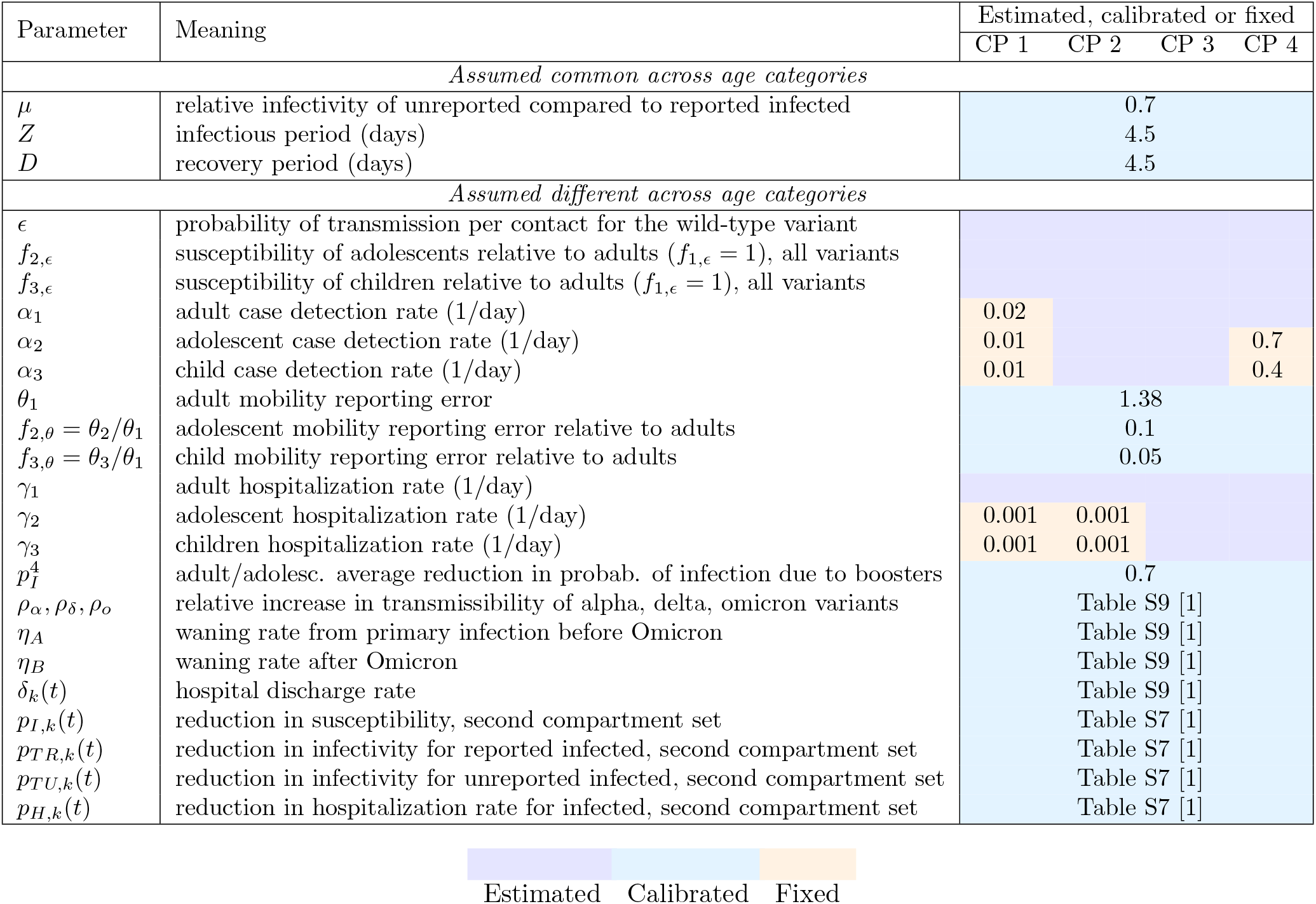
Summary of the main model parameters. This table provides an overview of the main model parameters as defined in the Methods. Parameters are classified as *estimated* : inferred from data, *calibrated* : values taken from [1] and other relevant studies, or *fixed* : set via grid search. CP: Counterfactual period

**Figure S5.**
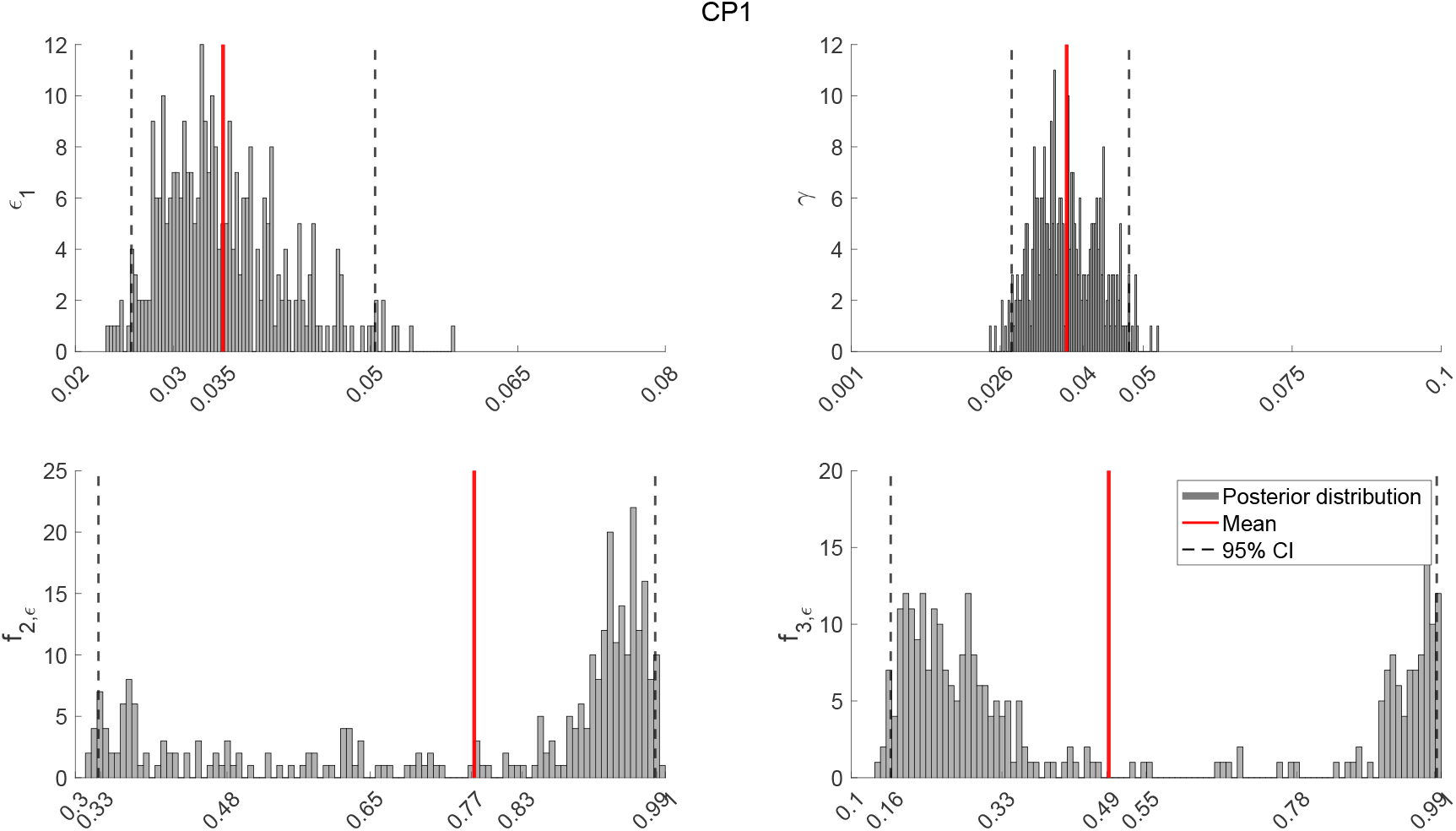
Parameter posterior distributions at the end of the period versus prior range - CP1. Posterior distributions at the end of the period compared to the prior range. Solid vertical red lines indicate posterior means; dashed vertical lines show 95% credible intervals (2.5%–97.5%). The x-axis corresponds to the prior range for each parameter.

**Figure S6.**
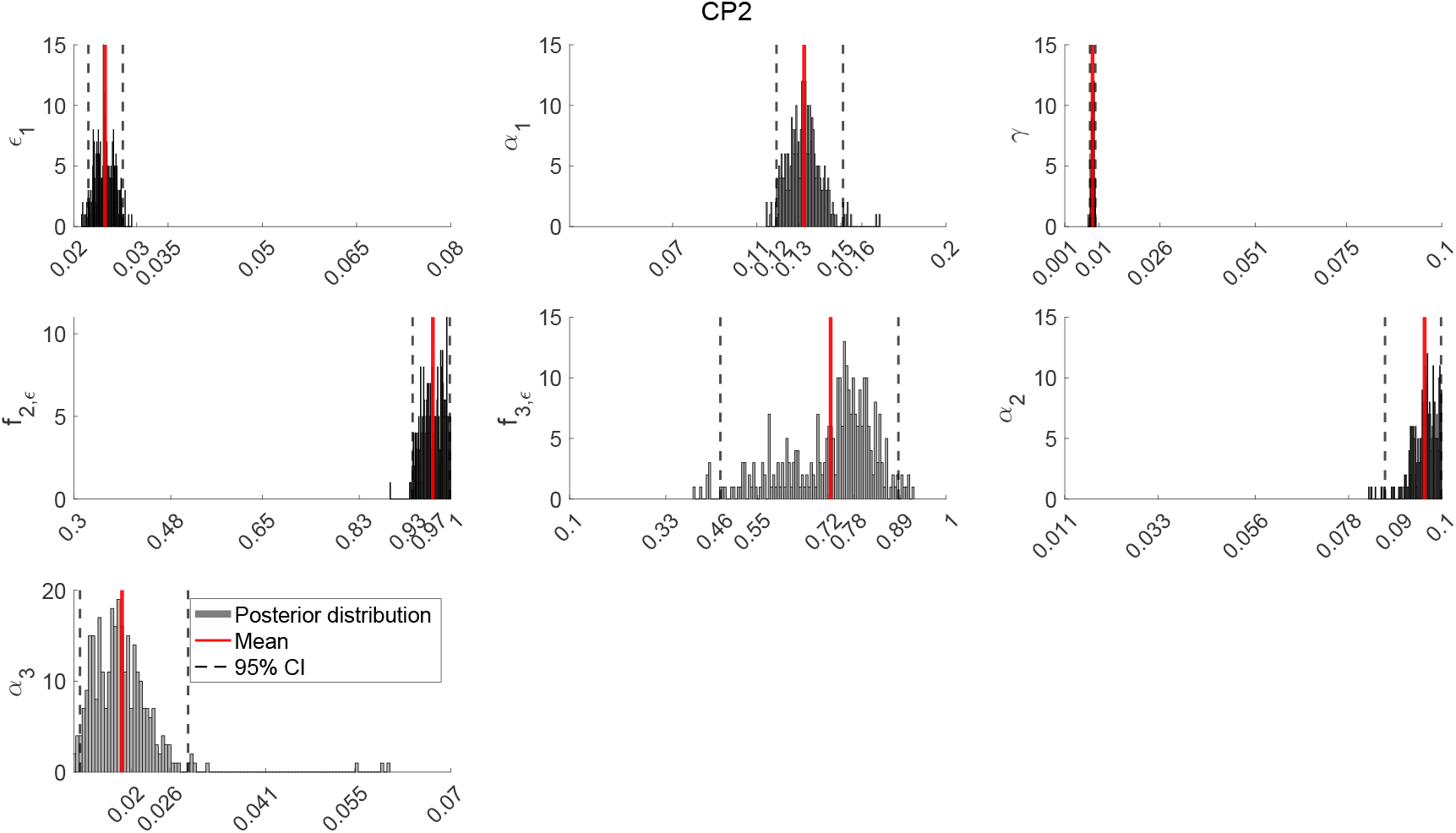
Parameter posterior distributions at the end of the period versus prior range - CP2. Posterior distributions at the end of the period compared to the prior range. Solid vertical red lines indicate posterior means; dashed vertical lines show 95% credible intervals (2.5%–97.5%). The x-axis corresponds to the prior range for each parameter.

**Figure S7.**
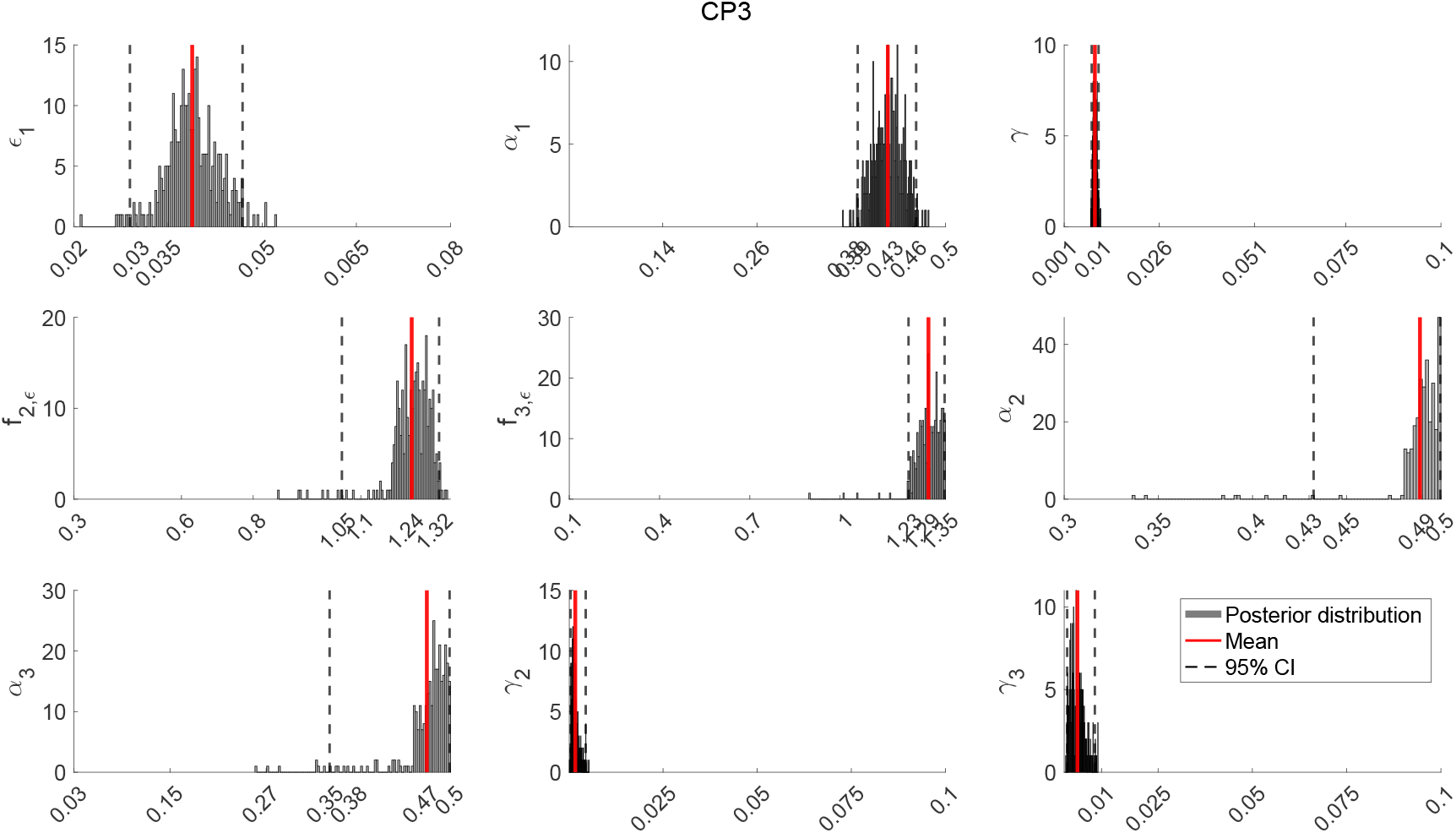
Parameter posterior distributions at the end of the period versus prior range - CP3. Posterior distributions at the end of the period compared to the prior range. Solid vertical red lines indicate posterior means; dashed vertical lines show 95% credible intervals (2.5%–97.5%). The x-axis corresponds to the prior range for each parameter.

**Figure S8.**
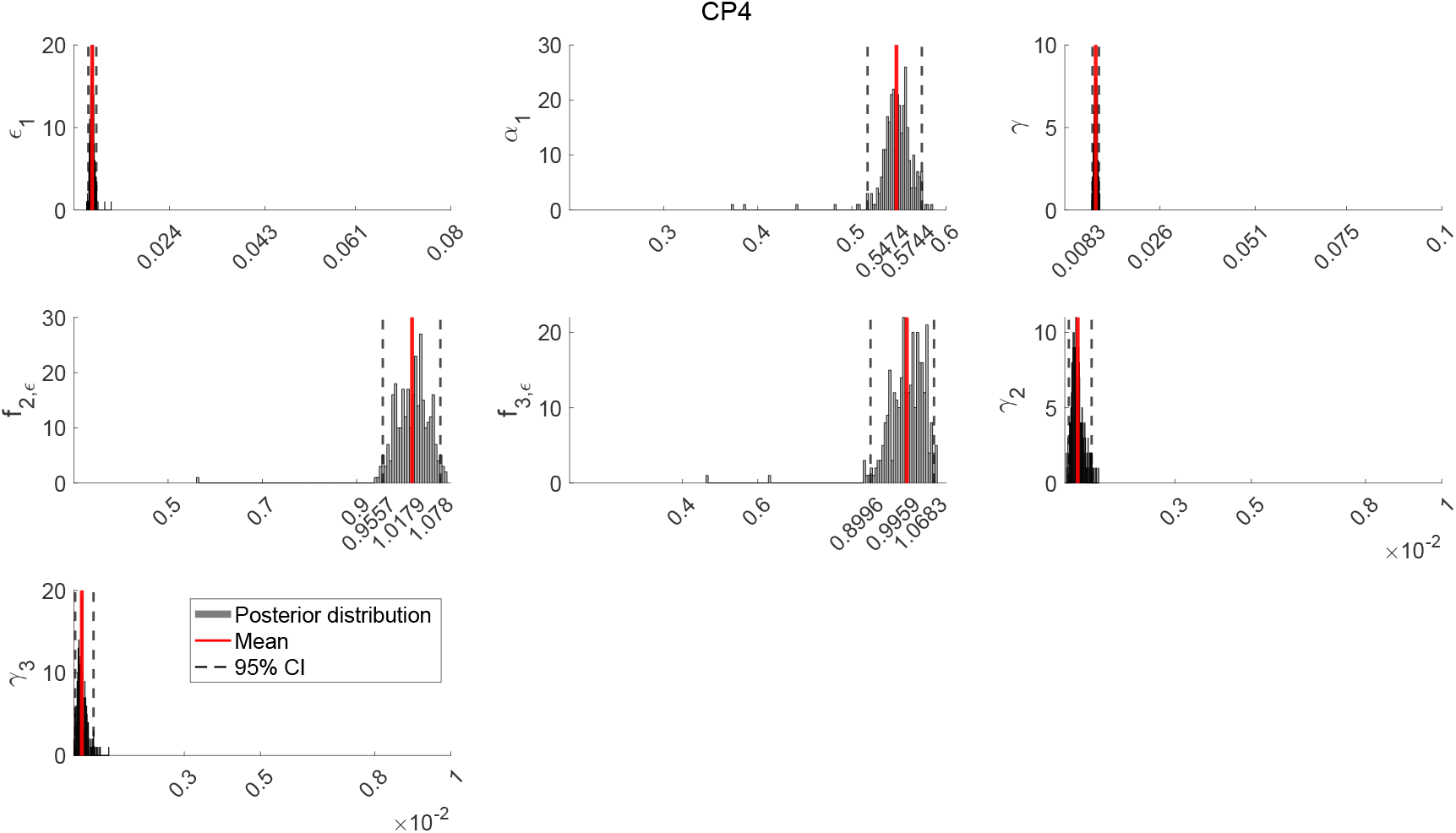
Parameter posterior distributions at the end of the period versus prior range - CP4. Posterior distributions at the end of the period compared to the prior range. Solid vertical red lines indicate posterior means; dashed vertical lines show 95% credible intervals (2.5%–97.5%). The x-axis corresponds to the prior range for each parameter.

### S4 Model fit and baseline counterfactual versus data

**Figure S9.**
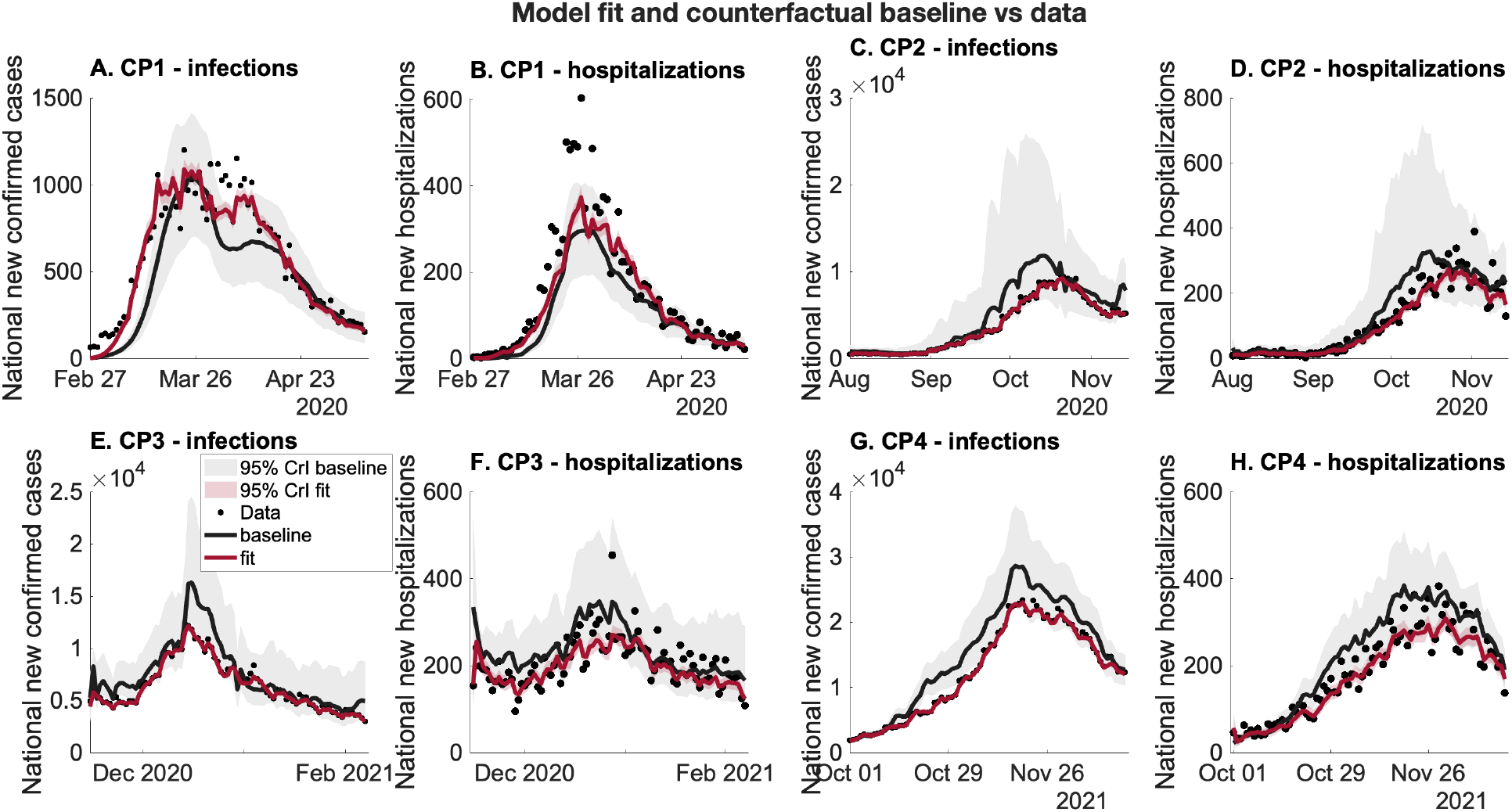
Model fit and baseline counterfactual compared with observed data. Time series of (**A**-**D**) national new confirmed cases and (**E**-**H**) new hospitalizations during the four considered periods (CP1–CP4). Black dots represent observed data, the red line shows the model fit, and the black line shows the baseline counterfactual scenario. Shaded areas indicate the 95% credible intervals (CrI) for the baseline (light gray) and the fit (light red). The x-axis shows time, and the y-axis shows daily national counts. A broader time window was considered for the fitting to ensure parameter identifiability. The model fit and baseline counterfactual reproduce the trends in the data.

### S5 Derivation of the next generation matrix

Based on the metapopulation in Equation S1, we now consider the following age-structured model in Equations S2. Compared to [1], we eliminated region-dependence and merged the populations to represent national dynamics between ages.

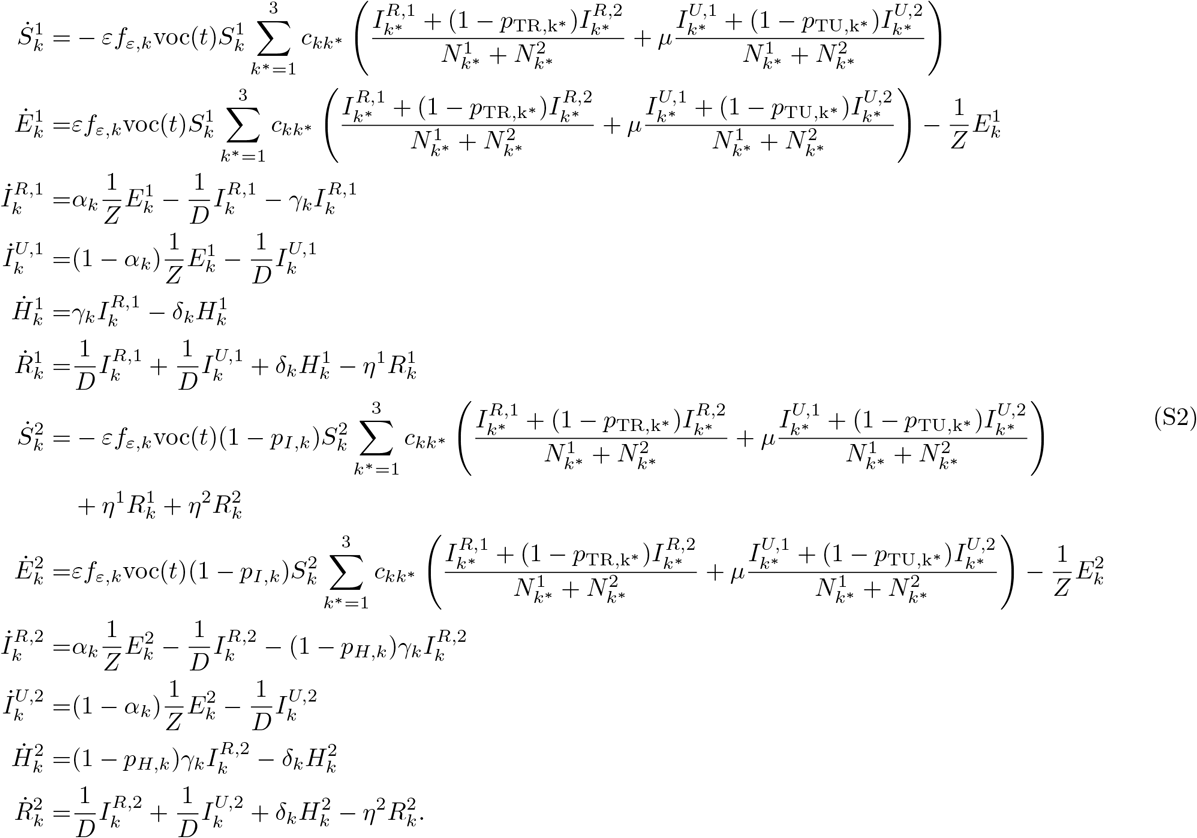

Similar indices from the original model are adapted, i.e., *a* represents the two sets of compartments, where *a* = 1 represents dynamics for naïve infections and *a* = 2 represents dynamics for breakthrough infections and reinfections, and *k* pertains to the age groups. With the twelve equations above (two for each compartment sets in *a*), the total number of equations is 36 for the three age groups in *k*. As compared to the 5184 equations of the metapopulation model, the calculation of the next-generation matrix for this simplified model is easier to analyze and compute.

#### S5.1 Via linear algebraic methods

Recall that the infection subsystem will consist of compartments 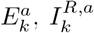, and 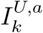. Using Equations S2,we get the following transition matrix **Σ** = [Σ_*k*_] which is a block diagonal matrix with elements

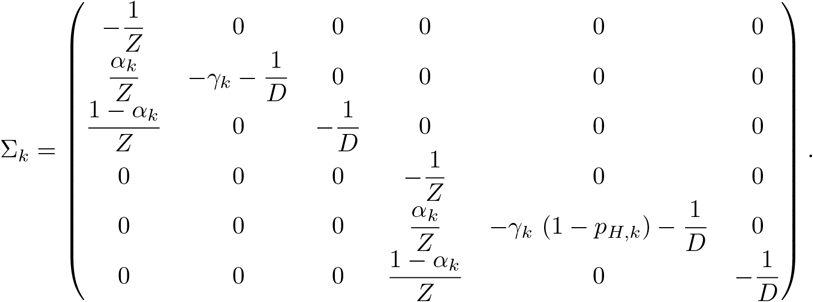

On the other hand, the transmission matrix **T** = [*T*_*ij*_] is

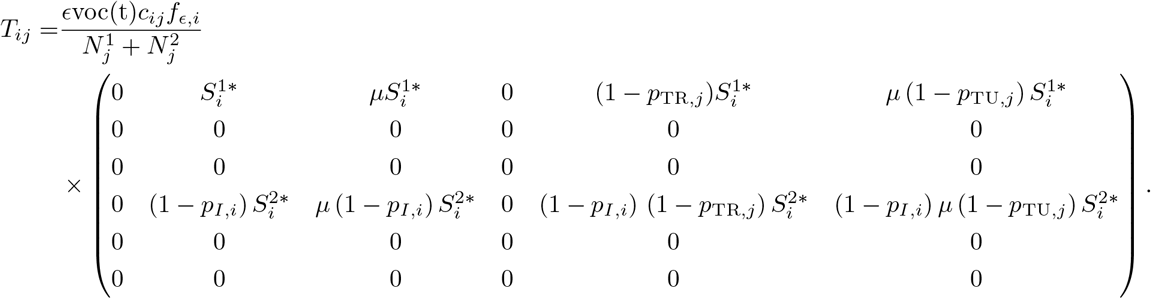

Both matrices are 18 ×18, corresponding to all possible infection states. From here, we get **K**_L_ = −**TΣ**^−1^ = [*K*_L,*ij*_], where

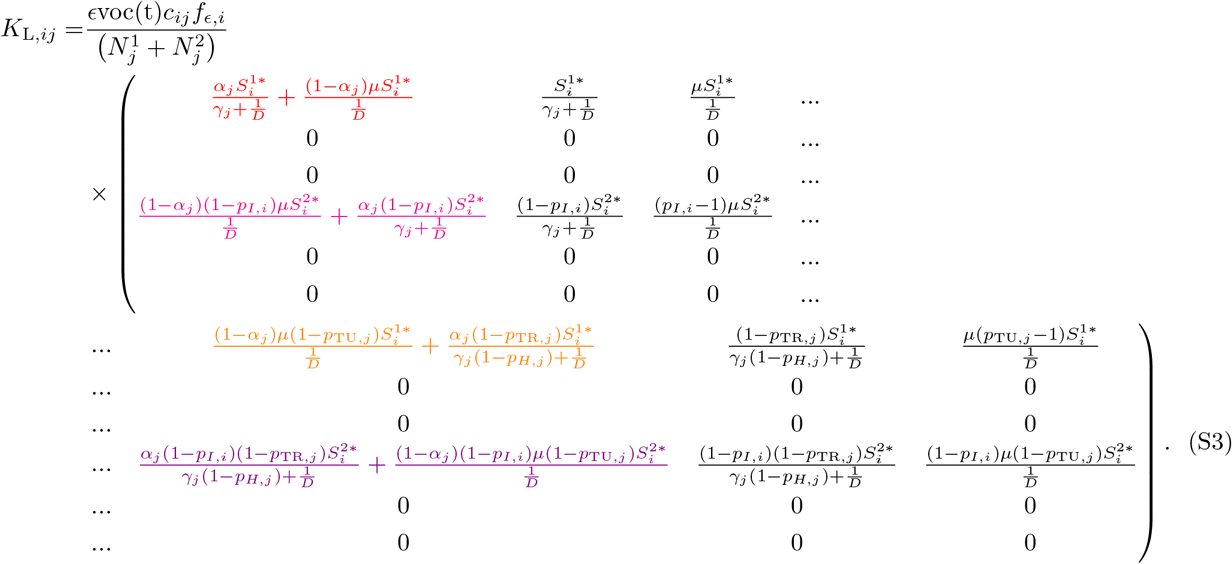

Corresponding to the indices for the states-at-infection, the auxiliary matrix **E** is

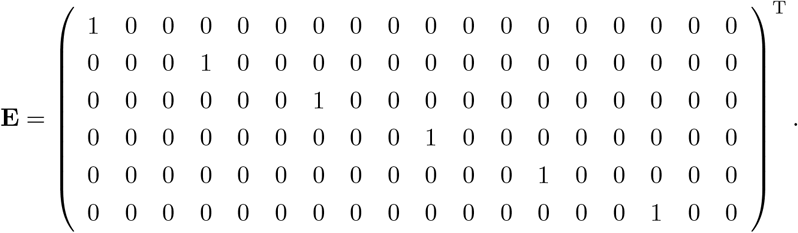

Hence, the next generation matrix is **K** = −**E**^T^ **TΣ**^−1^ **E** = [*K*_*mn*_] where

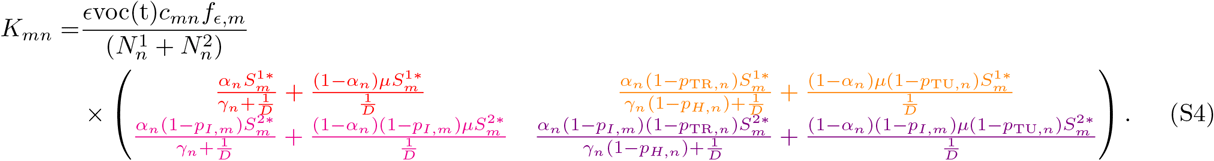

Note that the change of indices from (*i, j*) to (*m, n*) refers to the reduction of the matrix size from 18 × 18 for **K**_L_ to 6 × 6 for **K**.

#### S5.2 Via an epidemiological framework

Consider now the age-structured model in Equations S2. The states-of-infections are 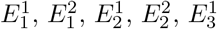, and 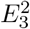 while the twelve states-on-infectiousness are 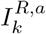, and 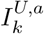 for *k* = 1, 2, 3 and *a* = 1, 2. For example, an individual in compartment 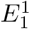 has a probability of 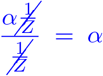 to progress to compartment 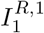. From here, the infected person becomes infectious and can infect an individual in 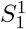 at a rate 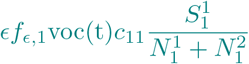 for a period 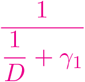.

Multiplying these two terms, we get the first entry of the next generation matrix. The other entry corresponds to the progression from 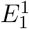 to 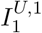 infecting 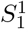 i.e.,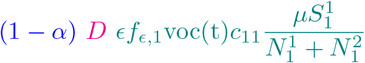. Hence, the first term is

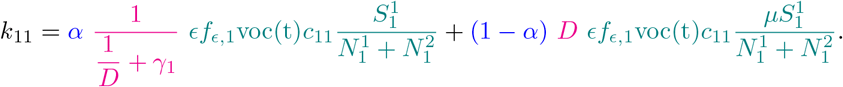

From here, a 6 × 6 matrix rewritten as a 3 × 3 block matrix in Equation S5 is created, i.e.

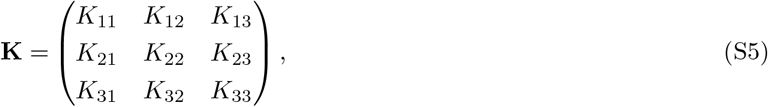

where *K*_*mn*_ is a 2 × 2 matrix of the transmission from the *n*th age group to the *m*th age group, detailed in Table S3.

**Table S3.** Defining terms inside the next generation matrix for the Dutch model using epidemiological recipe. The tabulated expressions are directly the entries of the submatrices in Equation S5. The transmission dynamics corresponding to the expressions are also shown.

| Element | Dynamics | Terms |
| --- | --- | --- |
| $k_{mn}^{11}$ | $E_n^1 \rightarrow I_n^{R,1} \dashrightarrow S_m^1$ | $\alpha_n \times \frac{1}{\frac{1}{D} + \gamma_n} \times \frac{\epsilon \text{voc}(t) f_{\epsilon,m} c_{mn} S_m^{1*}}{(N_n^1 + N_n^2)}$ |
| | $E_n^1 \rightarrow I_n^{U,1} \dashrightarrow S_m^1$ | $(1 - \alpha_n) \times D \times \frac{\epsilon \text{voc}(t) f_{\epsilon,m} c_{mn} \mu S_m^{1*}}{(N_n^1 + N_n^2)}$ |
| $k_{mn}^{12}$ | $E_n^2 \rightarrow I_n^{R,2} \dashrightarrow S_m^1$ | $\alpha_n \times \frac{1}{\frac{1}{D} + (1 - p_{H,n})\gamma_n} \times \frac{\epsilon \text{voc}(t) f_{\epsilon,m} c_{mn} (1 - p_{\text{TR},n}) S_m^{1*}}{(N_n^1 + N_n^2)}$ |
| | $E_n^2 \rightarrow I_n^{U,2} \dashrightarrow S_m^1$ | $(1 - \alpha_n) \times D \times \frac{\epsilon \text{voc}(t) f_{\epsilon,m} c_{mn} \mu (1 - p_{\text{TU},n}) S_m^{1*}}{(N_n^1 + N_n^2)}$ |
| $k_{mn}^{21}$ | $E_n^1 \rightarrow I_n^{R,1} \dashrightarrow S_m^2$ | $\alpha_n \times \frac{1}{\frac{1}{D} + \gamma_n} \times \frac{\epsilon \text{voc}(t) f_{\epsilon,m} c_{mn} (1 - p_{\text{I},m}) S_m^{2*}}{(N_n^1 + N_n^2)}$ |
| | $E_n^1 \rightarrow I_n^{U,1} \dashrightarrow S_m^2$ | $(1 - \alpha_n) \times D \times \frac{\epsilon \text{voc}(t) f_{\epsilon,m} c_{mn} (1 - p_{\text{I},m}) \mu S_m^{2*}}{(N_n^1 + N_n^2)}$ |
| $k_{mn}^{22}$ | $E_n^2 \rightarrow I_n^{R,2} \dashrightarrow S_m^2$ | $\alpha_n \times \frac{1}{\frac{1}{D} + (1 - p_{H,n})\gamma_n} \times \frac{\epsilon \text{voc}(t) f_{\epsilon,m} c_{mn} (1 - p_{\text{I},m}) (1 - p_{\text{TR},n}) S_m^{2*}}{(N_n^1 + N_n^2)}$ |
| | $E_n^2 \rightarrow I_n^{U,2} \dashrightarrow S_m^2$ | $(1 - \alpha_n) \times D \times \frac{\epsilon \text{voc}(t) f_{\epsilon,m} c_{mn} (1 - p_{\text{I},m}) \mu (1 - p_{\text{TU},n}) S_m^{2*}}{(N_n^1 + N_n^2)}$ |

Table S3 lists the terms for each submatrix *K*_*mn*_. Thus, the matrix *K*_*mn*_ can be written as

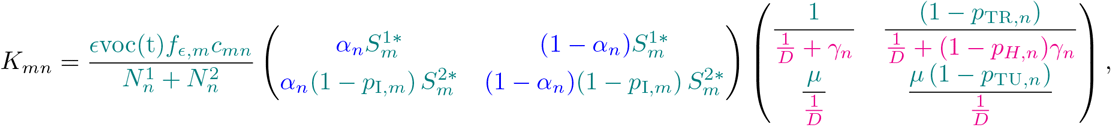

which we can readily programmed to get the time-varying reproduction number of the system. This equation is in agreement with the obtained next-generation matrix using the linear algebraic methods in Equations S4.

### S6 Calculation of cumulative elasticities

For indices *m* and *n* of the six population groups combining age (adults, adolescents, and children) and immune classes (naïve and partially immune), we calculate the sensitivity, *s*_*mn*_(*t*), representing the rate of change of ℛ_*e*_(*t*) with respect to the (*m, n*)-th entry of the next generation matrix, **K** for time *t*. Each term is simply the partial derivative 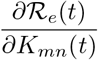. To construct the sensitivity matrix *S*(*t*) = [*s*_*mn*_(*t*) (*t*)], Caswell [2] derived the expression for the element *s*_*mn*_(*t*) as,

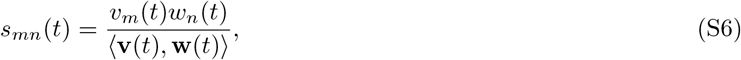

where *v*_*m*_(*t*) is the *m*-th entry of the left eigenvector **v**(*t*), *w*_*n*_(*t*) is the *n*-th entry of the right eigenvector **w**(*t*), and ⟨·, ·⟩ denotes the dot product.

The elasticity matrix, E(*t*) = [*e*_*mn*_(*t*)], is a scaled version of S(*t*), with its elements expressed as

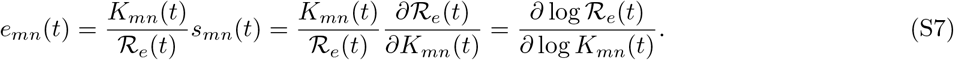

In matrix notation, the elasticity can be written as,

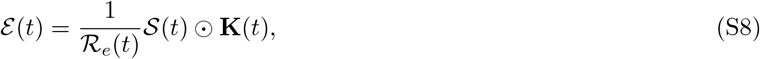

where ⊙ refers to the element-wise product. Each element of the elasticity matrix, ℰ_*mn*_(*t*), represents the contribution of the corresponding *K*_*mn*_(*t*) entry to ℛ_*e*_(*t*). From here, the cumulative elasticity can be obtained by summing along the column of the matrix, i.e.

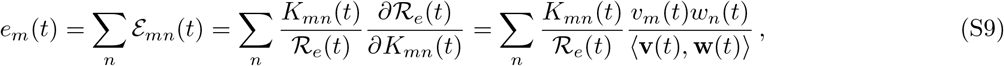

where **v**(*t*) and **w**(*t*) are the left and right eigenvectors of *K*_*mn*_(*t*) associated with the largest eigenvalue ℛ_*e*_(*t*), and ⟨·, ·⟩ denotes the dot product.

### S7 Time-varying cumulative elasticities

**Figure S10.**
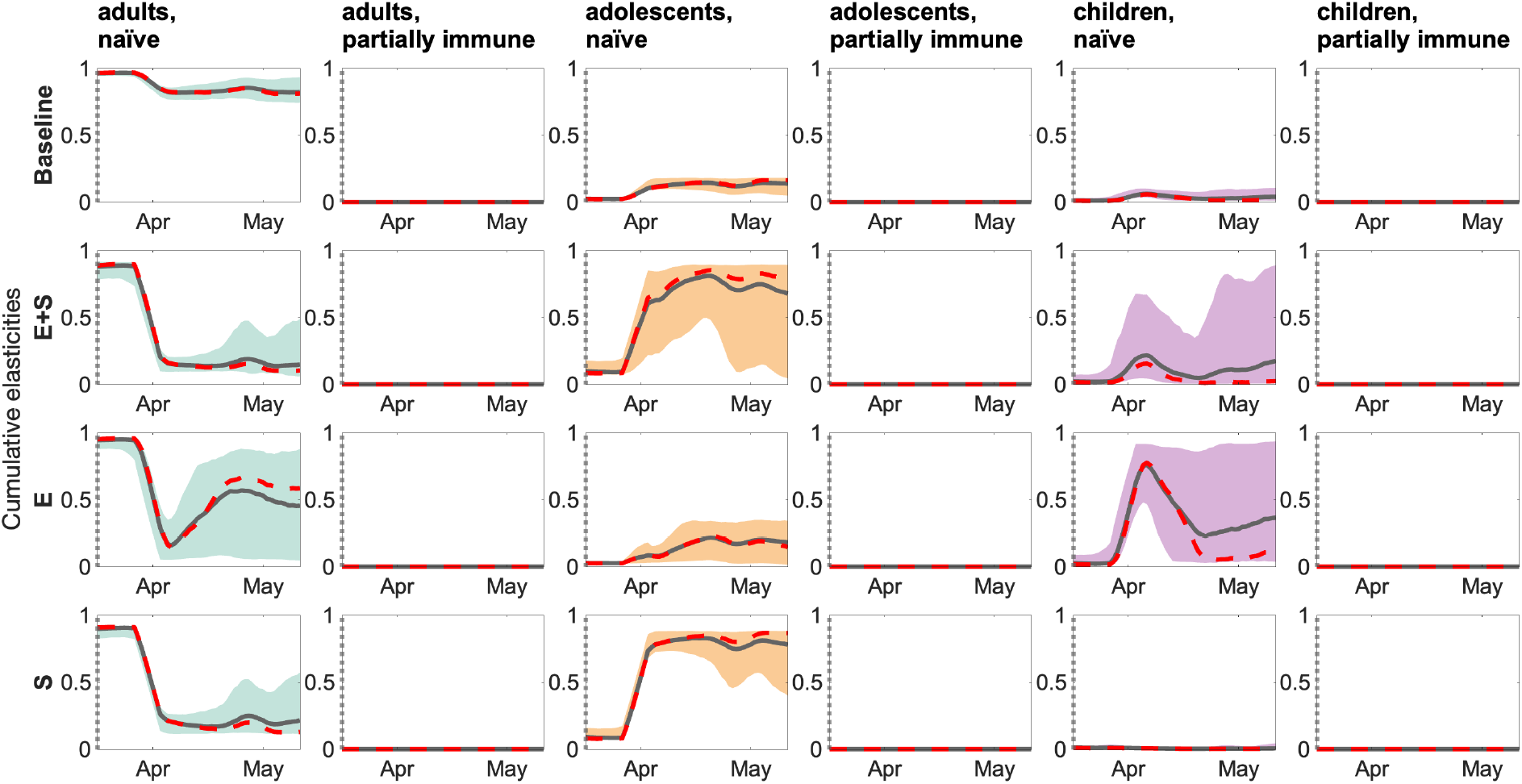
Cumulative elasticities by age group and scenario – CP1. The gray solid lines are the mean trajectories, whereas the red dashed lines are the median trajectories. The gray dotted vertical lines indicate the start of the intervention protocols. Filled areas represent the 95% credible intervals.

**Figure S11.**
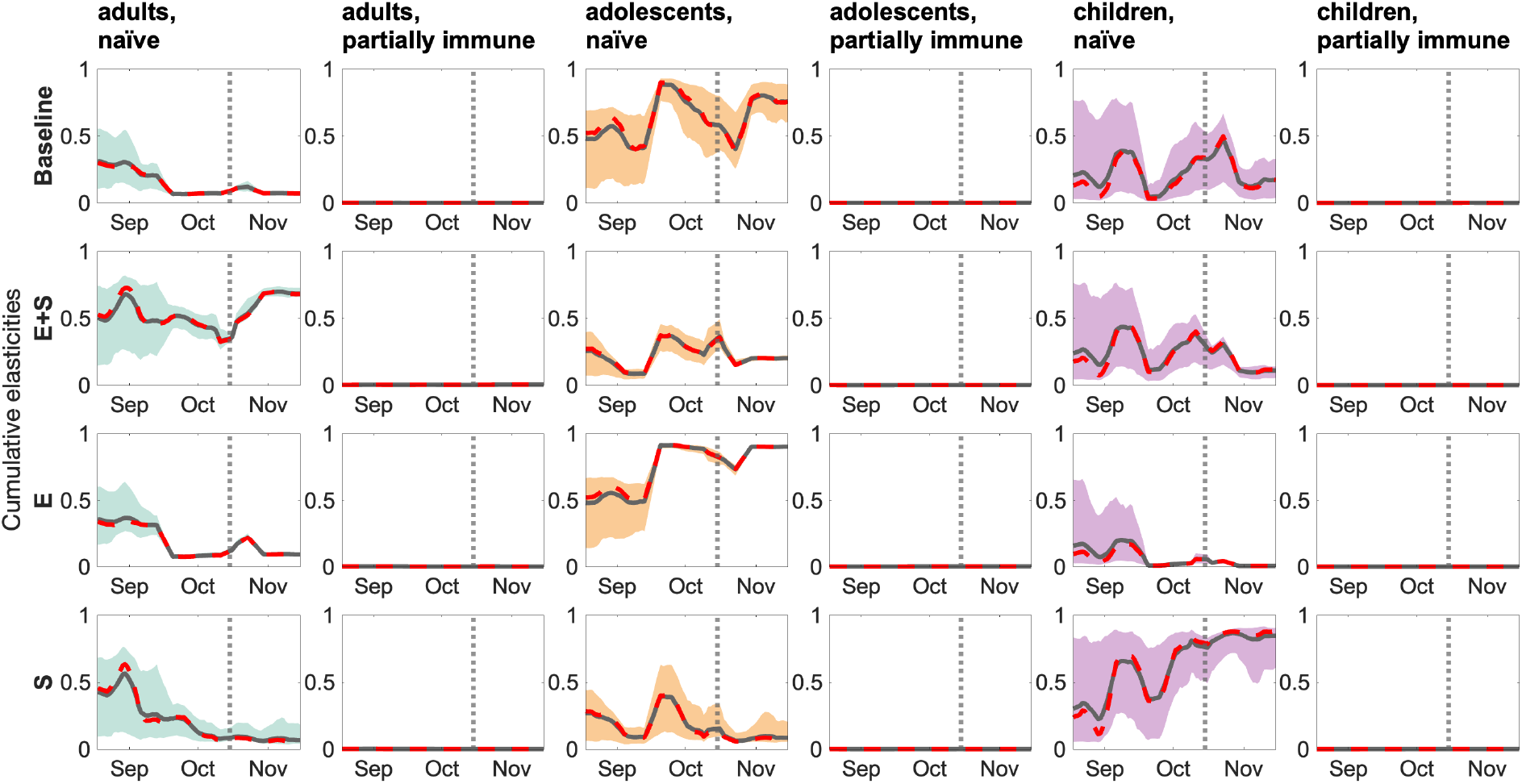
Cumulative elasticities by age group and scenario – CP2. The gray solid lines are the mean trajectories, whereas the red dashed lines are the median trajectories. The gray dotted vertical lines indicate the start of the intervention protocols. Filled areas represent the 95% credible intervals.

**Figure S12.**
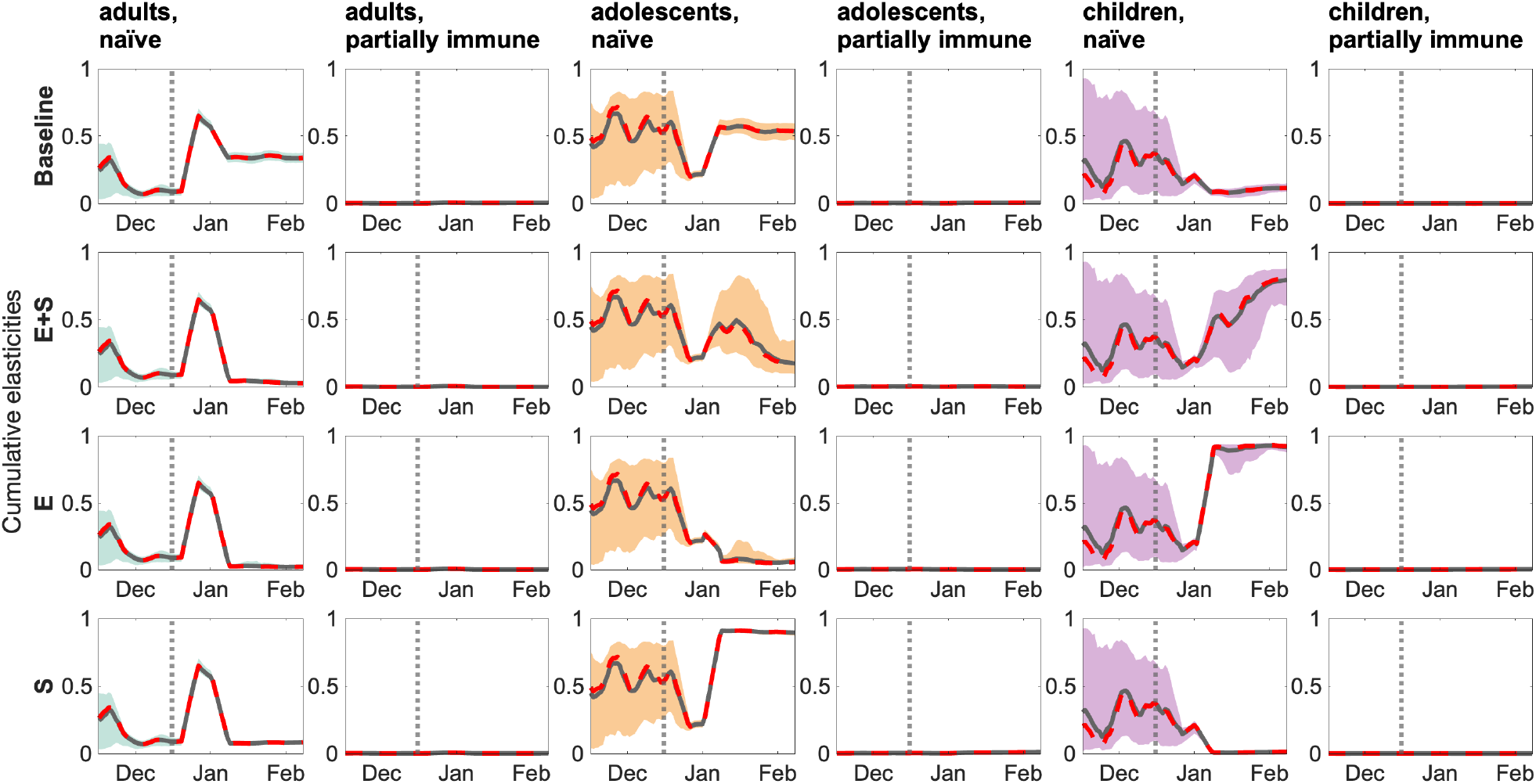
Cumulative elasticities by age group and scenario – CP3. The gray solid lines are the mean trajectories, whereas the red dashed lines are the median trajectories. The gray dotted vertical lines indicate the start of the intervention protocols. Filled areas represent the 95% credible intervals.

**Figure S13.**
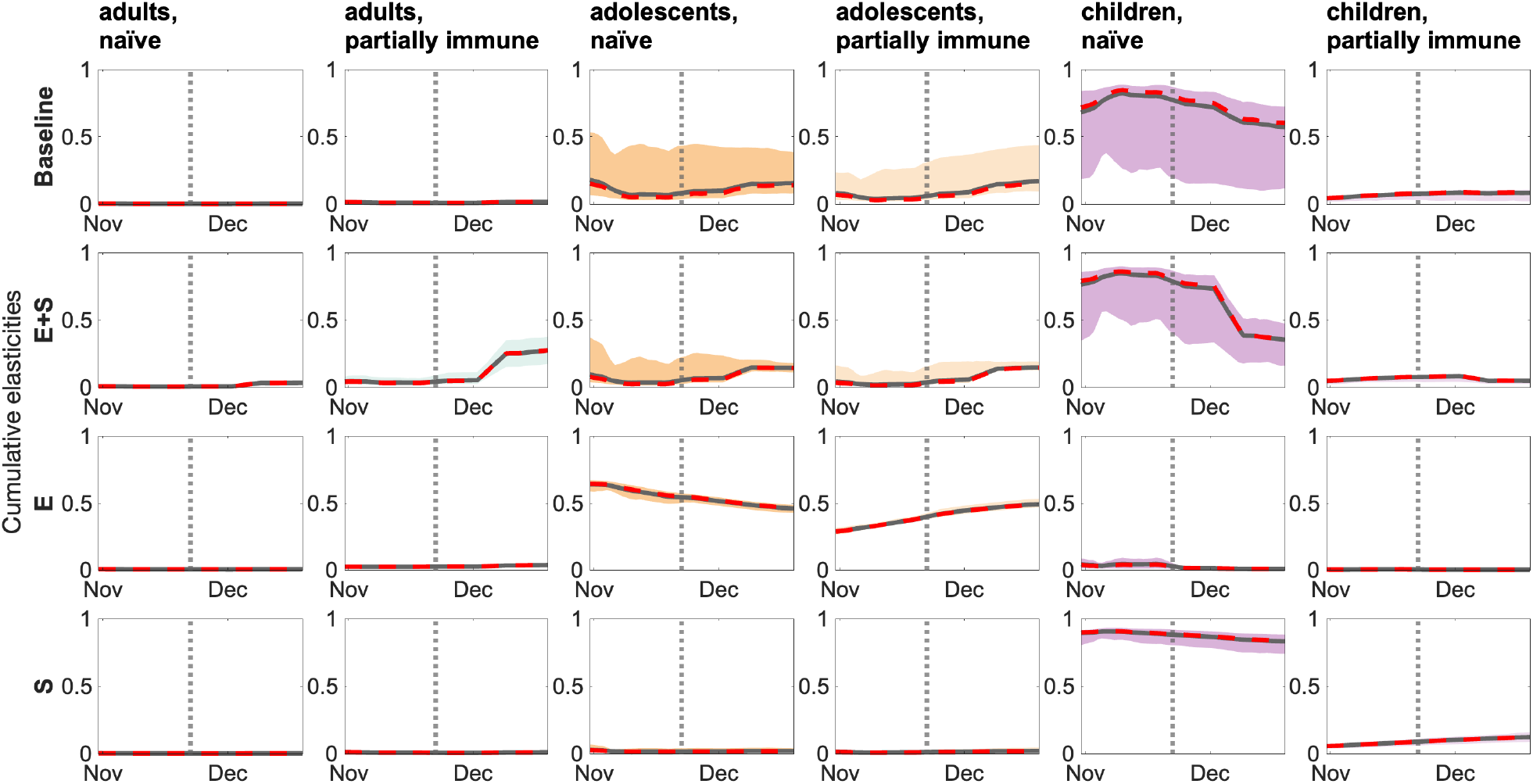
Cumulative elasticities by age group and scenario – CP4. The gray solid lines are the mean trajectories, whereas the red dashed lines are the median trajectories. The gray dotted vertical lines indicate the start of the intervention protocols. Filled areas represent the 95% credible intervals.

### S8 Proportion of immune and susceptible populations

**Figure S14.**
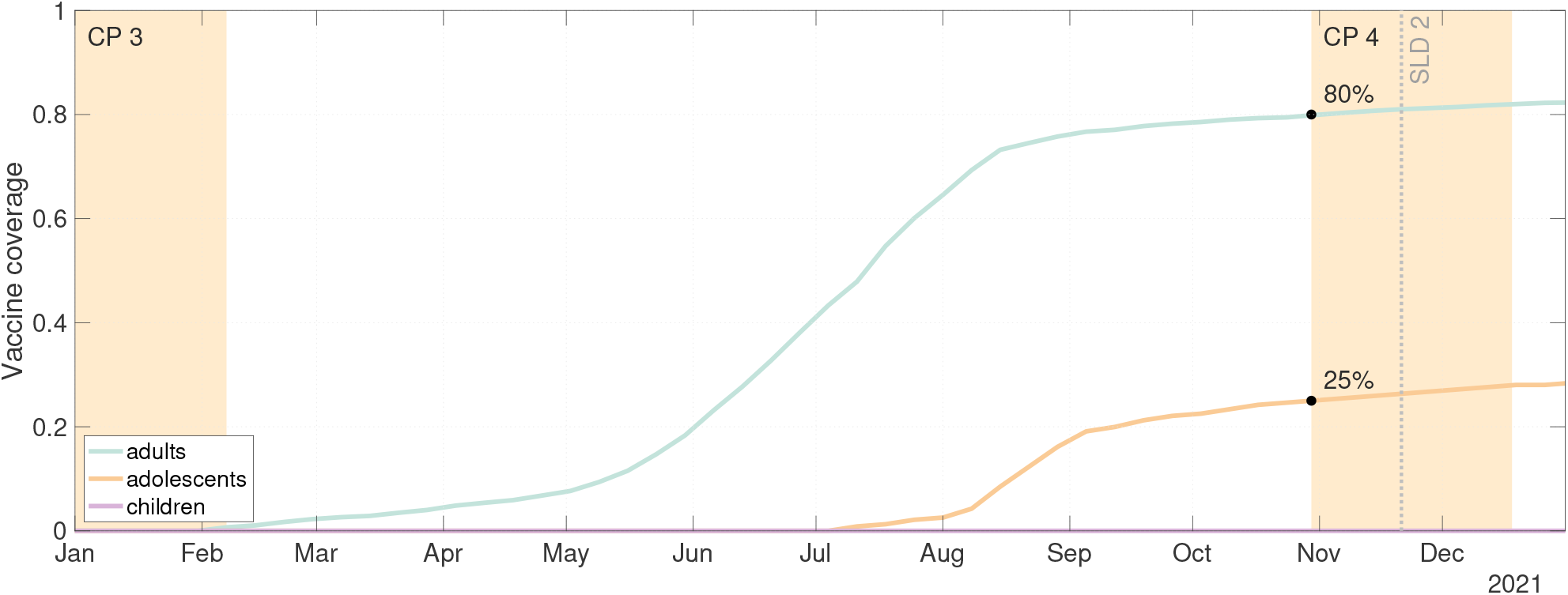
Vaccination coverage in 2021 from the RIVM dashboard. Age-stratified proportion of the Dutch population that has been vaccinated for the year 2021. Shaded areas highlight the counterfactual periods we analyze in 2021 — CP3 and CP4 — which are time windows we defined around the true date of specific measures implementation. The timing of the second semi-lockdown is indicated by the vertical dashed gray line annotated with SLD 2. CP: counterfactual period. LD: lockdown. SLD: semi-lockdown.

**Figure S15.**
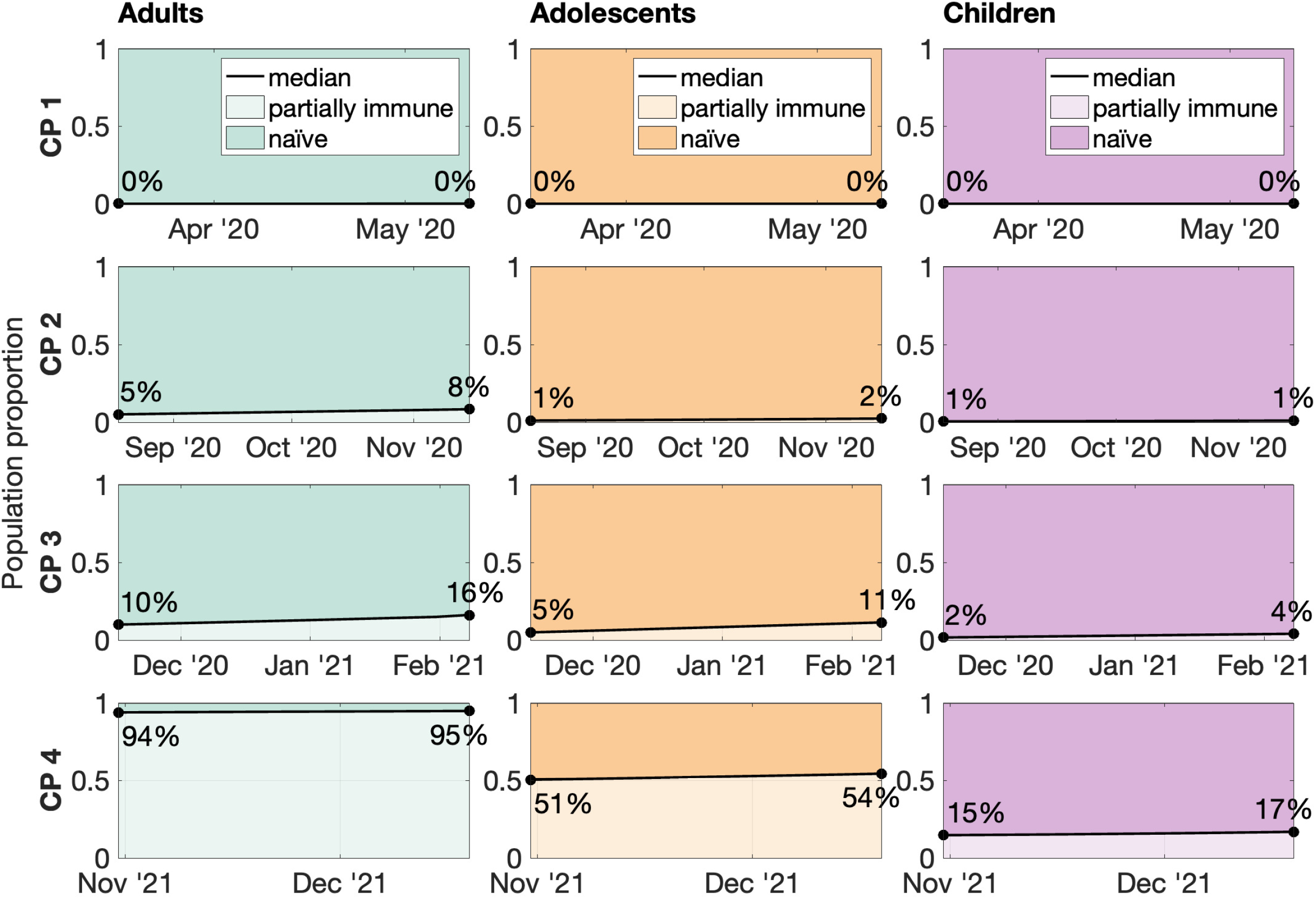
Proportion of naïve and partially immune population. Estimated proportion of individuals classified as naïve (*a* = 1) or partially immune (*a* = 2) in adults, adolescents, and children for all periods considered. Shaded areas indicate the proportion of the population that is partially immune (light colored) or naïve (colored). Annotations with black filled circles refer to median values for partially immune proportions at the start and end of each period refer. Solid black lines represent the median estimates. CP: counterfactual period.

**Figure S16.**
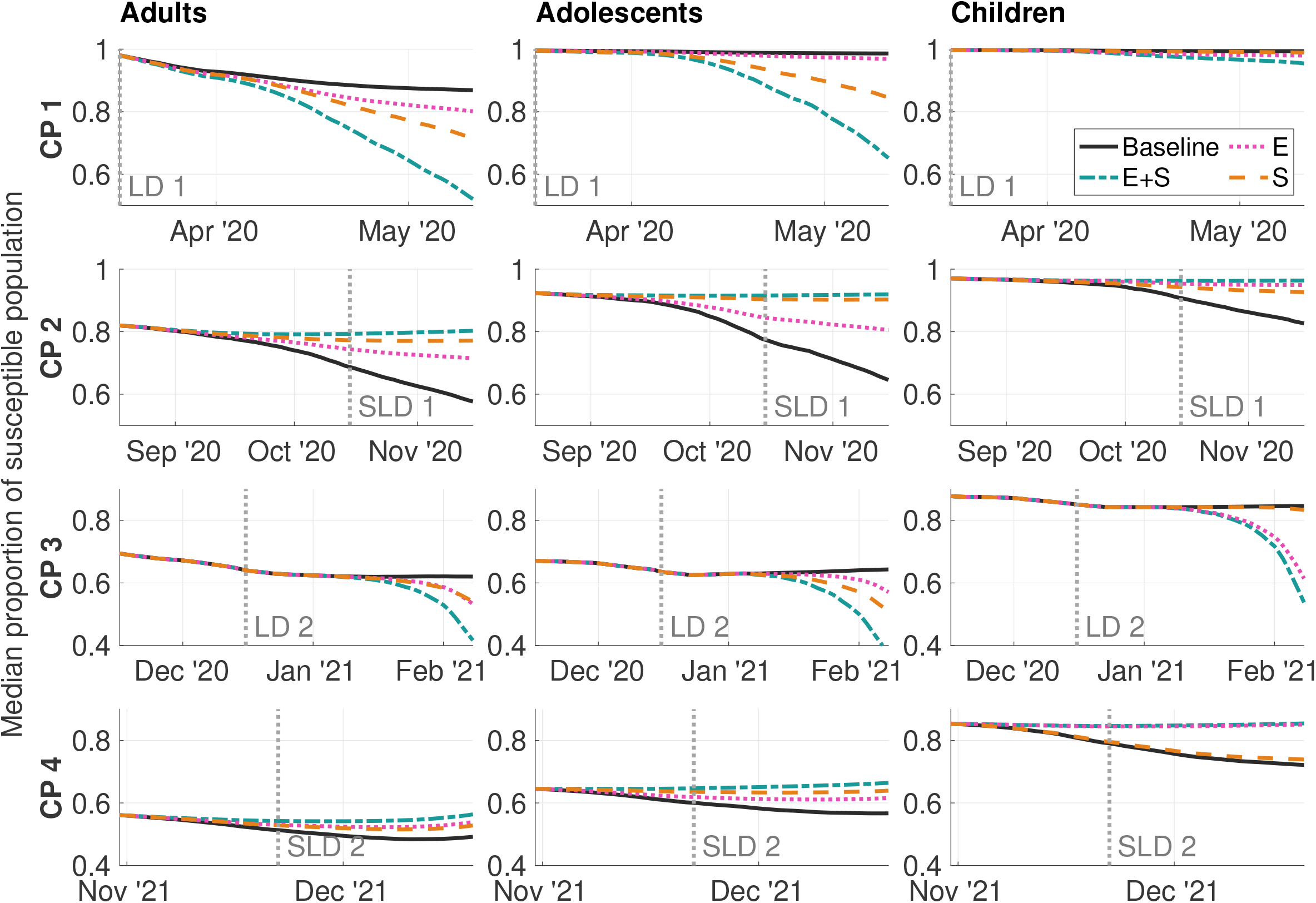
Proportion of susceptible population over time. Estimated proportion of susceptible (naïve + partially susceptible) individuals among adults, adolescents, and children for all periods considered under the baseline and three counterfactual scenarios. The baseline is shown as a black solid line. Counterfactual scenarios include measures for both elementary and secondary schools (E+S, blue green dashed-dotted), measures for elementary schools only (E, pink dotted), and measures for secondary schools only (S, orange dashed). The vertical dotted lines indicate the implementation of the lockdowns and semi-lockdowns corresponding to the counterfactual periods. CP: counterfactual period. LD: lockdown. SLD: semi-lockdown.

### S9 Impact of counterfactual scenarios on disease burden

**Table S4.**
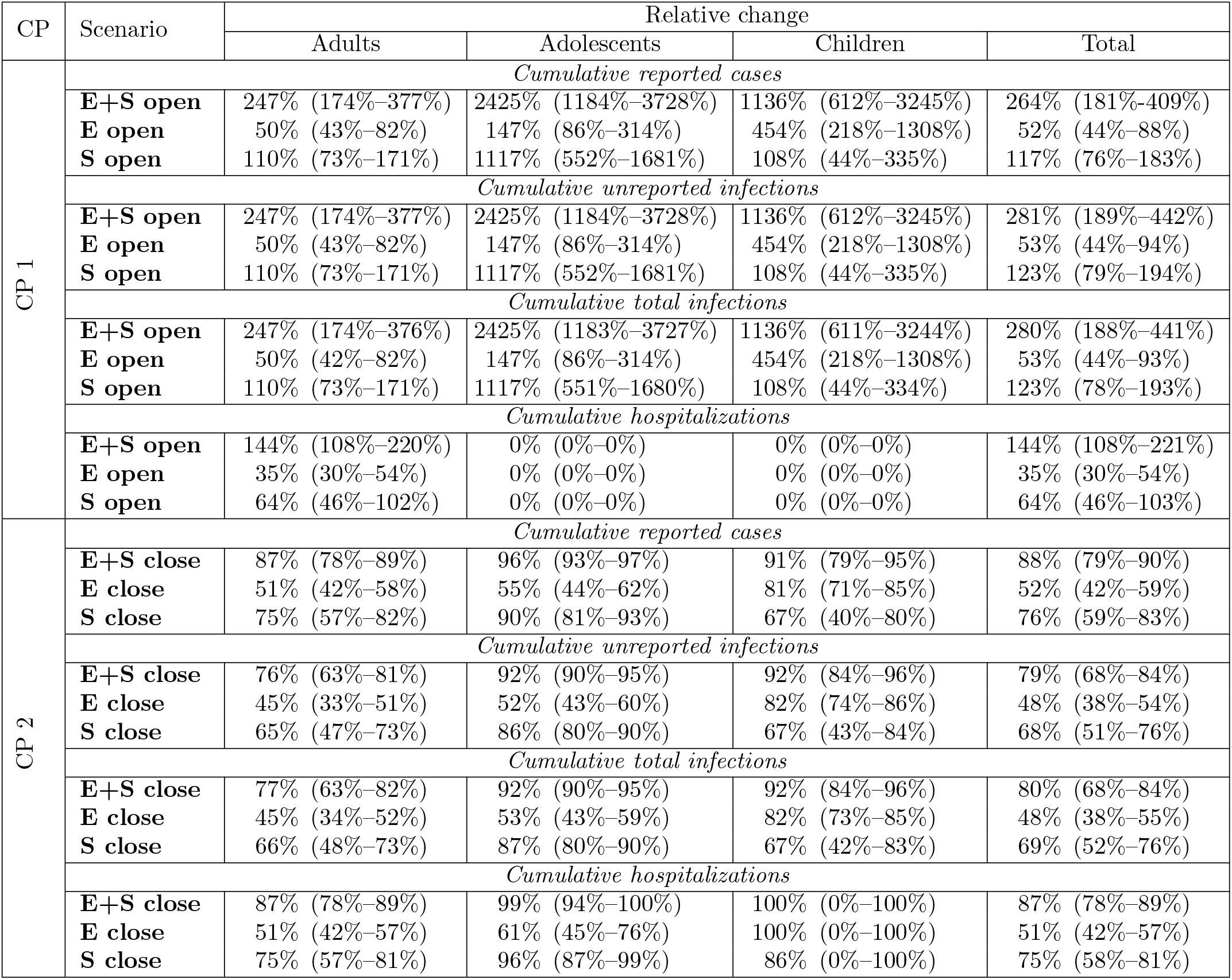
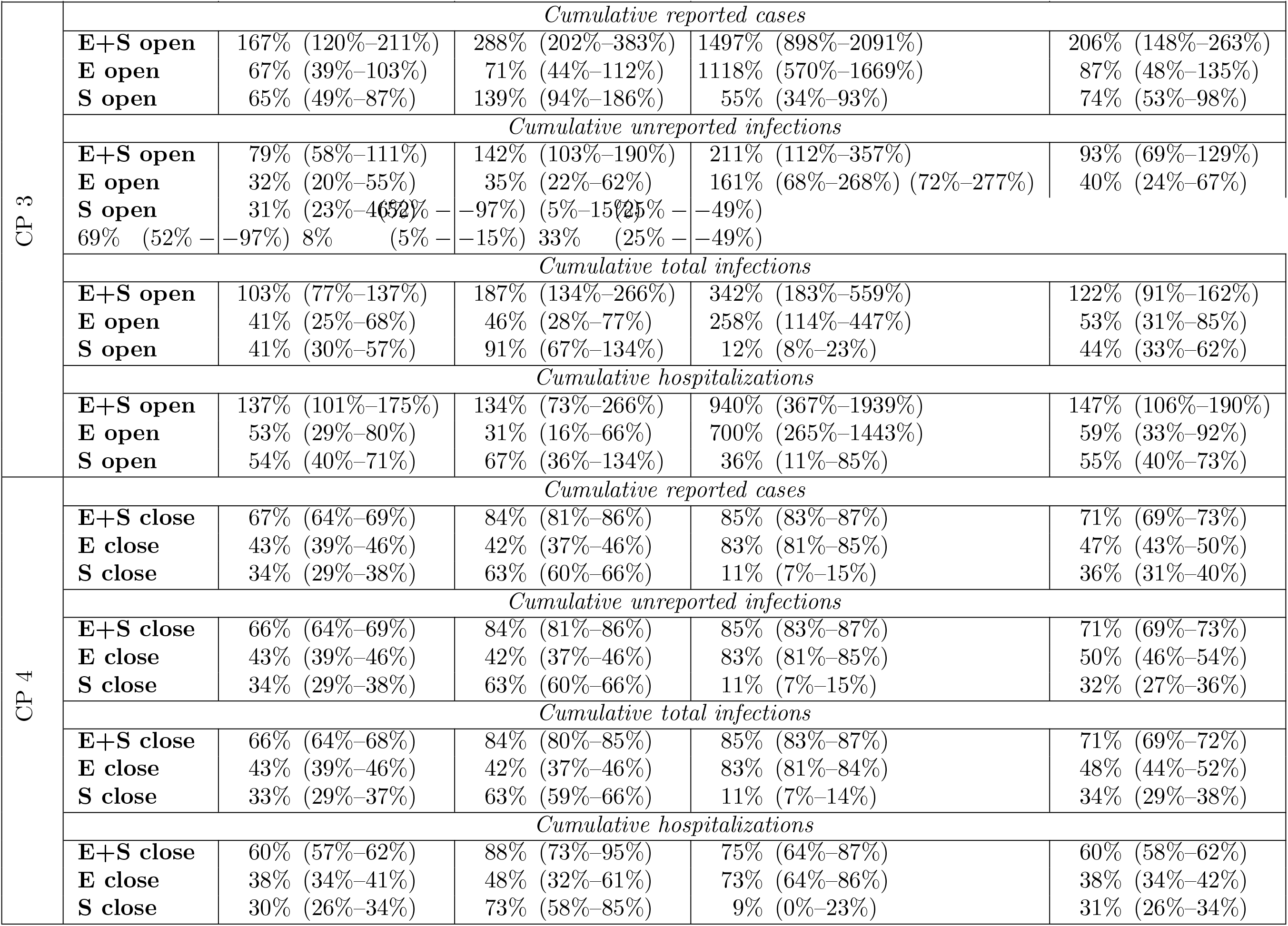
Period-wise relative changes in disease burden with respect to the baseline. Relative increase (CP1) and relative decrease (CP2) of cumulative infections and hospitalizations at the end of the period for the entire population and three age groups separately. Median values are shown alongside 95% credible intervals. Period-wise relative changes of disease burden with respect to the baseline. Relative increase (CP3) and relative decrease (CP4) of cumulative infections and hospitalizations at the end of the period for the entire population and three age groups separately. Median values are shown alongside 95% credible intervals.

